# Decomposing polygenic risk of coronary artery disease into distinct biological processes

**DOI:** 10.64898/2026.10.08.26365088

**Authors:** Jakub Morze, Kirk Smith, Marcin Kierczak, Göran Bergström, Hui Li, Anders Gummesson, Tove Fall, Stefan Söderberg, Tino Ebbers, J. Gustav Smith, Rikard Landberg, Paul Franks, Tuuli Lappalainen, Miriam Udler, Clemens Wittenbecher

## Abstract

Standard polygenic risk scores capture overall genetic liability to coronary artery disease (CAD) but do not reveal the biological processes contributing to risk in an individual. Here, we applied data-driven latent factor decomposition to 526 genome-wide significant CAD-associated variants, using their associations with 78 CAD-related phenotypic traits. We identified eight genetic components corresponding to cholesterol-enriched lipoproteins, triglyceride-rich lipoproteins, lipoprotein(a), metabolically unhealthy normal weight, obesity, endothelial dysfunction, coagulation and cardiac remodeling. Process-specific polygenic risk scores (pPRSs) captured biological profiles beyond those reflected by conventional risk factors. In UK Biobank (n = 458,905), the Swedish CArdioPulmonary bioImage Study (n = 24,904) and Mass General Brigham Biobank (n = 53,848), pPRSs were consistently associated with clinical and subclinical CAD and showed distinct biomarker, proteomic and comorbidity profiles. These findings establish a framework for biologically resolved genetic CAD risk and may inform process-specific approaches to precision prevention.

## Main

Coronary artery disease (CAD) is the leading cause of death worldwide.^1^ An individual’s CAD risk is determined by the interplay of biological processes, including disturbances in lipid metabolism, body fat accumulation and distribution, and tissue-specific immune and remodeling processes. The current paradigm for CAD prevention rests on identifying individuals at high risk of CAD and prioritizing them for risk-mitigating interventions.^2,3^

Genetic factors make a substantial contribution to CAD susceptibility, with twin studies estimating heritability at approximately 40–60% and common genetic variants accounting for up to approximately half of this heritable liability.^4,5^ Over the last two decades, genome-wide association studies (GWAS) have identified several hundred independent loci associated with CAD across more than a million participants of diverse ancestries.^6–8^ Polygenic risk scores aggregate the effects of these variants into a summary metric of inherited risk and identify high-risk individuals more accurately than scores based on conventional clinical factors alone.^9^ Hence, genetic risk assessment has emerged as a potential tool for refining stratified CAD prevention.

The pleiotropic genetic CAD liability reflects the diversity of biological processes involved in CAD development.^10^ The current aggregated CAD polygenic risk scores identify who is at elevated risk but do not indicate which biological processes are responsible for high CAD susceptibility in each individual. This information, however, may be critical for next-generation precision prevention approaches that aim to tailor interventions according to the individual biological risk profile. In the context of other complex diseases, data-driven latent factor models have been applied to decompose the genetic architecture into central biological processes. Previous studies in type 2 diabetes (T2D), metabolic dysfunction-associated steatotic liver disease (MASLD) and obesity have derived process-specific polygenic risk scores (pPRSs) associated with distinct phenotypic profiles and differential disease incidence and complication trajectories.^11–16^ A data-driven approach to decompose overall genetic liability for CAD into central biological processes and quantify inherited risk in each process separately is currently lacking.

Here, we performed latent component decomposition of genome-wide significant CAD risk variants based on their association profiles across a broad panel of CAD-relevant phenotypic traits. Using Bayesian non-negative matrix factorization (bNMF), we derived eight latent genetic components corresponding to distinct biological processes acting in different tissue compartments. In three large cohort studies spanning more than 500,000 individuals, pPRSs showed distinct and biologically plausible association patterns with molecular risk factors, imaging-defined subclinical atherosclerosis, clinical coronary events, and comorbidity patterns. We further tested whether process-specific genetic liability modified associations between established, modifiable risk factors and incident myocardial infarction, suggesting candidate subgroups in which risk-factor modification may be most consequential. Together, these analyses establish a biologically resolved framework for CAD genetic liability and identify testable hypotheses for process-specific precision prevention.

## Results

### Latent genetic components of CAD risk map to distinct biological processes

To extract latent genetic components underlying CAD risk, we adapted a high-throughput bNMF pipeline previously applied to T2D.^11–13^ Using genome-wide significant CAD risk variants and their associations with a curated panel of disease-relevant traits as input, the pipeline decomposes variant-trait association structure into latent components (**Fig. 1**). Single variants can carry continuous weights across multiple components.

**Figure 1.**
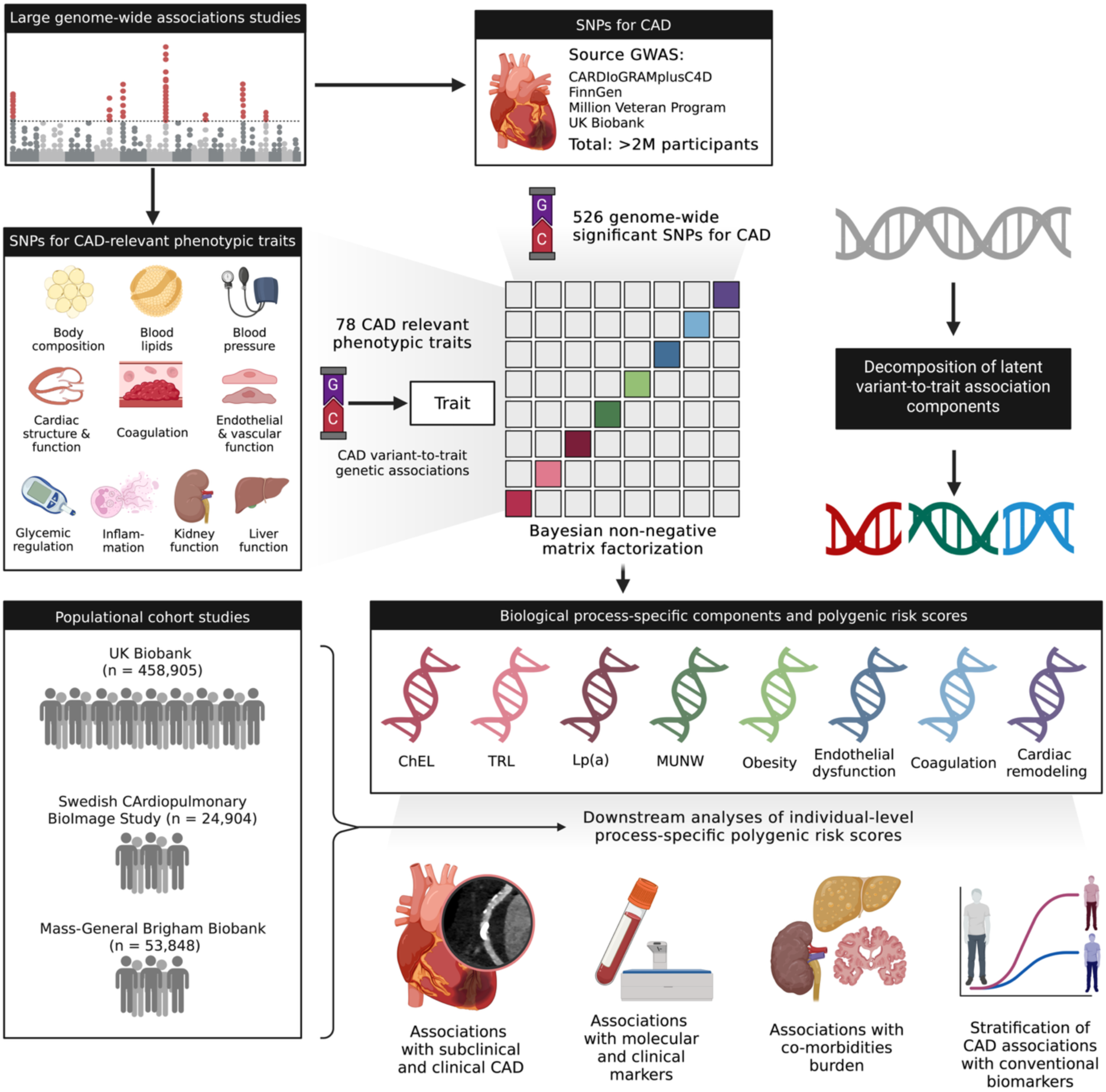
Conceptual overview of the analytical framework. Abbreviations: CAD, coronary artery disease; ChEL, cholesterol-enriched lipoproteins; Lp(a), lipoprotein(a); MUNW, metabolically unhealthy normal weight; SNP, single-nucleotide polymorphism; TRL, triglyceride-rich lipoproteins.

We assembled summary statistics from seven CAD GWAS spanning >2M participants of predominantly European ancestry (**Supplementary Table 1**), together with the largest available GWAS for each of 87 manually curated phenotypic traits relevant to CAD pathophysiology (**Supplementary Table 2**). After variant and trait quality control, 526 independent CAD risk variants (**Supplementary Table 3**) and 78 traits were used for latent structure learning.

Across 50 independent runs, the bNMF CAD risk variant-to-trait analysis converged on eight stable latent genetic components (**Fig. 2a**, **Supplementary Tables 4**−**5**). We refined previous pipeline versions by substituting a single global threshold^11–13^ with component-specific selection thresholds for top-weighted genetic variants, which yielded more balanced components. Comparing the global threshold with the component-specific one, the median number of variants per component increased from 11 to 33 while preserving variant-to-component specificity (**Extended Data Fig. 1**). A convenient implementation of the updated bNMF pipeline is publicly available (see Code availability).

**Figure 2.**
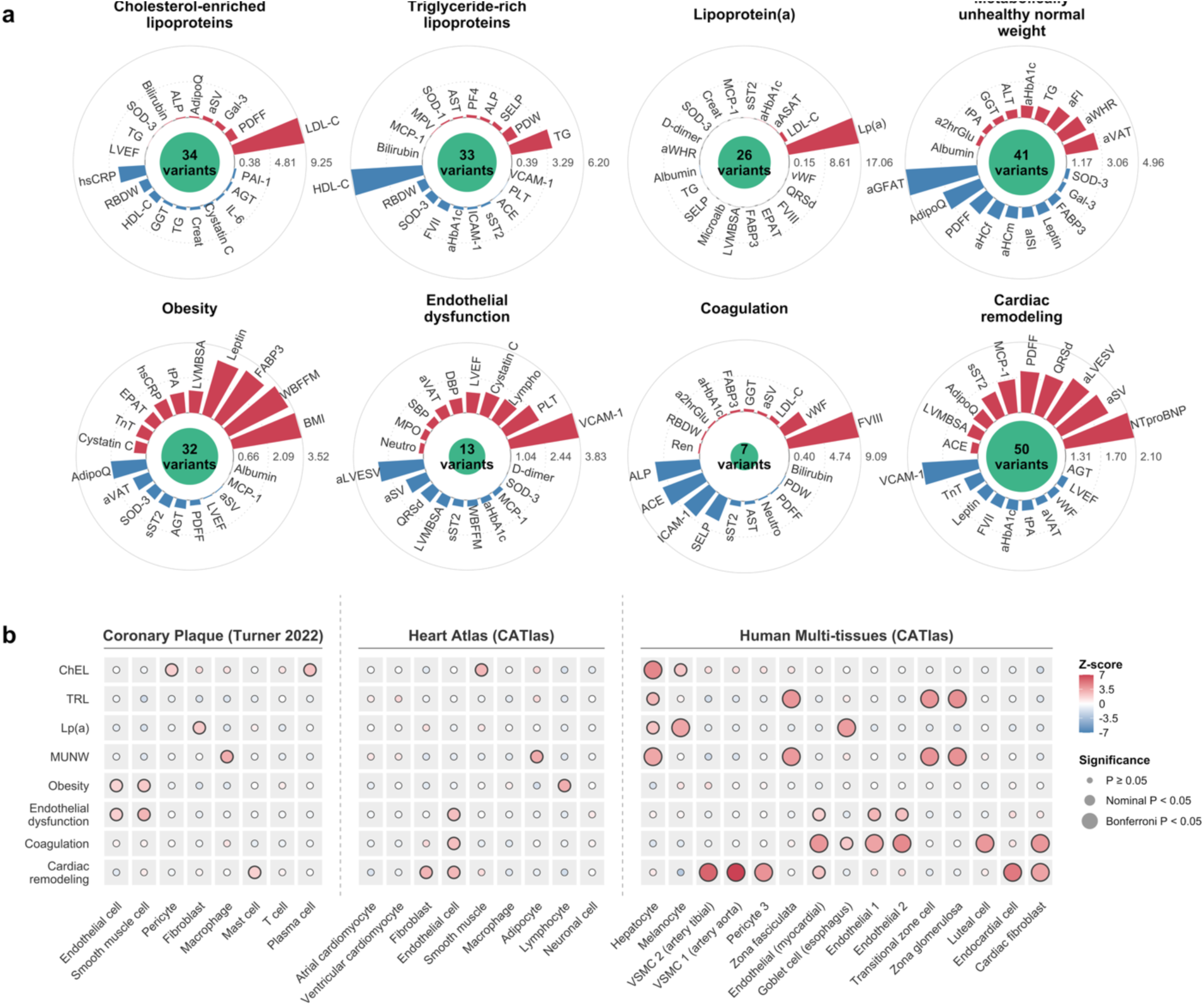
Composition of eight latent genetic components of CAD risk and enrichment in cell-type-resolved accessible chromatin. (a) Radial bar plots show the top-weighted positive (red) and top-weighted negative (blue) traits for each of the eight CAD genetic components identified by Bayesian non-negative matrix factorization (bNMF). The top 10 traits in each direction are shown per component, ranked by absolute component weight; bar length is proportional to the component weight. The central circle is sized proportionally to the number of variants assigned to each component (range 7–50). Trait abbreviations are summarized in Supplementary Table 2. (b) Dot matrix summarizing g-chromVAR enrichment of component-defining variants in cell-type-specific accessible chromatin across three epigenomic reference atlases: a single-cell ATAC-seq atlas of human coronary atherosclerotic plaque (8 cell types), a single-cell ATAC-seq atlas of the adult human heart (9 cell types), and the whole-body CATLAS reference of candidate cis-regulatory elements (111 adult cells; only cells reaching P_Bonferroni_<0.05 are shown). Dot color encodes the enrichment z-score. Dot size denotes statistical significance: non-significant (P ≥ 0.05), nominal (P < 0.05), and multiple-testing corrected (P_Bonferroni_ < 0.05, applied per component across 128 cell types).

To interpret and label each component, we examined its top-weighted variants and traits (**Table 1**). Three of the eight latent genetic components mapped onto the metabolism of apoB-containing lipoproteins. The ***Cholesterol-enriched lipoproteins*** (ChEL) component (34 variants) was defined by low-density lipoprotein (LDL) cholesterol, with top-weighted loci (*APOE*, *APOC1*, *LDLR*, *PSRC1*, *HNF1A*) converging on hepatic LDL receptor-mediated clearance.^17,18^ The ***Triglyceride-rich lipoproteins*** (TRL) component (33 variants) was defined by high positive weights for triglycerides (TG) and negative weights for high-density lipoprotein (HDL) cholesterol, with top-weighted loci (*CETP*, *LPL*, *ANGPTL4*, *SLC18A1*, *GALNT2*, *TRIB1*) encoding key regulators of TRL metabolism.^19^

**Table 1.** Characteristics of eight genetic components of coronary artery disease risk and their hypothesized mechanisms.

| Component | Key traits | Key loci | Expected physiological impact |
| --- | --- | --- | --- |
| Cholesterol-enriched lipoproteins (34 variants) | LDL-C (+) | *APOE, APOC1, LDLR, PSRC1, HNF1A* | Hypercholesterolemia |
| Triglyceride-rich lipoproteins (33 variants) | TG (+) | *CETP, LPL, SLC18A1, ANGPTL4, GALNT2, TRIB1* | Hypertriglyceridemia |
| Lipoprotein(a) (26 variants) | Lp(a) (+) | *LPA, SLC22A2, PLG* | Elevated Lp(a) |
| Metabolically unhealthy normal weight (41 variants) | aVAT (+), aWHR (+), aFI (+), TG (+), aHbA1c (+) | *VEGFA, COBLL1, LOC646736 (IRS1), ZNF664-RFLNA, CCDC92, MAP3K1* | Insulin resistance with lipodystrophy-like fat distribution |
| Obesity (32 variants) | BMI (+), WBFFM (+), FABP3 (+), Leptin (+) | *FTO, MC4R, TMEM18* | Increased total adiposity |
| Endothelial dysfunction (13 variants) | VCAM-1 (+), PLT (+), Lympho (+), Cystatin C (+) | *SH2B3, CENPW, NOS3, HNF1A, SERPINA1, GUCY1A1* | Endothelial activation and vascular inflammation |
| Coagulation (7 variants) | FVIII (+), vWF (+) | *ABO, ST3GAL4* | Prothrombotic state |
| Cardiac remodeling (50 variants) | NTproBNP (+), aSV (+), aLVESV (+), QRSd (+) | *GOSR2, COG5, ABCA8, WWP2, ZNF318, MAP3K7CL* | Vascular and cardiac fibrosis |

Traits abbreviations are explained in Supplementary Table 2.

The **Lipoprotein(a)** [Lp(a)] component (26 variants) had Lp(a) as its top positive weight and was led by *LPA*, the principal genetic determinant of plasma Lp(a) concentrations,^20^ with *SLC22A2* and *PLG* in the surrounding 6q25.3–q26 region. Together, these components distinguish CAD-associated genetic signals linked to cholesterol-enriched lipoproteins, triglyceride-rich lipoproteins and Lp(a).

Two genetic components mapped onto the metabolic consequences of body fat distribution and quantity. The ***Metabolically unhealthy normal weight*** (MUNW) component (41 variants) was defined by positive loadings for body mass index (BMI)- adjusted markers of android fat, fasting insulin and glycated hemoglobin (HbA1c), and negative loadings for BMI-adjusted markers of gynoid fat, insulin sensitivity and adiponectin (AdipoQ). Top-weighted loci (*VEGFA*, *COBLL1*, *IRS1*, *ZNF664-RFLNA*, *CCDC92*, *MAP3K1*) encode established determinants of body fat distribution and insulin signaling.^21,22^ In contrast, the ***Obesity*** component (32 variants) loaded positively on BMI, fat-free mass, heart-type fatty acid-binding protein and leptin, and negatively on AdipoQ and BMI-adjusted visceral adipose tissue. Its top-weighted loci (*FTO*, *MC4R*, *TMEM18*) have been implicated in central appetite and energy-balance regulation.^23,24^ Opposing anthropometric loadings separate a lipodystrophy-like pattern of fat redistribution in MUNW from increased total adiposity in the Obesity component.

Two latent genetic components were linked to endothelial and hemostatic processes. The ***Endothelial dysfunction*** component (13 variants) loaded positively on vascular cell adhesion molecule 1 (VCAM-1) and platelet and lymphocyte counts. Its top-weighted loci include enzymes of nitric oxide signaling (*NOS3*, *GUCY1A1*)^25,26^ and regulators of vascular inflammation (*SH2B3*, *SERPINA1*).^27^ The ***Coagulation*** component (7 variants) was defined by positive loadings on coagulation factor VIII (FVIII) and von Willebrand factor (vWF), and negative loadings on alkaline phosphatase, intercellular adhesion molecule 1 and P-selectin. Its top-weighted loci (*ABO*, *ST3GAL4*) are related to glycosyltransferases regulating clearance of vWF and FVIII.^28^ Therefore, the data-driven latent structure learning approach yielded distinct genetic CAD risk axes for endothelial activation and coagulation factor availability.

The final component, ***Cardiac remodeling*** (50 variants), loaded positively on N-terminal pro-B-type natriuretic peptide (NTproBNP), left ventricular volumes and QRS duration, and negatively on VCAM-1 and troponin T. Its top-weighted loci map to cardiac development (*GOSR2*),^29^ transforming growth factor-β-driven fibrosis (*WWP2*)^30^ and heart failure susceptibility loci (*ZNF318*, *MAP3K7CL*)^31,32^. This component thus reflects an axis of unfavorable myocardial remodeling.

### Enrichment of component-defining variants in cell-type-resolved accessible chromatin

Because the latent genetic components were defined by variant–trait associations alone, we sought orthogonal support for their biological interpretation by testing whether component-associated variants were enriched in cell-type-resolved accessible chromatin. We used three atlases, spanning coronary atherosclerotic plaque, the human heart and multiple human tissues (**Fig. 2b**, **Supplementary Table 6**). After multiple-testing correction (P_Bonferroni_<0.05), ChEL-associated variants were enriched in hepatocyte-accessible chromatin, whereas TRL-associated variants were enriched in chromatin accessible in adrenal cortical cells of the zona glomerulosa, zona fasciculata and transitional zone. Lp(a)-associated variants were enriched in esophageal goblet cell- and melanocyte-accessible chromatin. MUNW-associated variants shared the adrenal cortical enrichment observed for TRL and were additionally enriched in hepatocyte-accessible chromatin. Coagulation-associated variants were enriched in endothelial cell-accessible chromatin, including general and myocardial endothelial cells, as well as in cardiac fibroblast- and luteal cell-accessible chromatin. Cardiac remodeling-associated variants showed the strongest enrichment overall, in vascular smooth muscle cells of the aorta and tibial artery, and were further enriched in endocardial cell-, pericyte- and cardiac fibroblast-accessible chromatin. At nominal significance (P<0.05), Endothelial dysfunction-associated variants were consistently enriched in endothelial cell-accessible chromatin across all three atlases, MUNW-associated variants in adipocytes from both the heart and multi-tissue atlases and in plaque macrophages, and Obesity-associated variants in heart-atlas lymphocytes and endocrine cell types (pancreatic alpha, gastric neuroendocrine and thyroid follicular cells). Hence, the biological processes inferred for each component from trait and loci composition were broadly corroborated by the cell-type specific chromatin accessibility enrichment analyses.

### Process-specific polygenic scores, subclinical CAD, and clinical coronary event risk

In the UK Biobank (UKBB; n=458,905), the Swedish CArdioPulmonary bioImage Study (SCAPIS; n=24,904) and the Mass General Brigham Biobank (MGBB; n=53,848), we constructed individual-level pPRS for each of the eight components by summing the individual component variants multiplied by their component-specific weights. Correlations between each pPRS and the total CAD polygenic risk score were modest (Spearman ρ = 0.13-0.45), while correlations among pPRS were low (|ρ| ≤ 0.14), indicating that the pPRSs capture distinct dimensions of genetic CAD liability (**Extended Data Fig. 2**).

First, we validated that each pPRS was consistently associated with clinical coronary events in all three cohorts (Cases: UKBB n=53,907; SCAPIS, n=705; MGBB, n=3250), comparing the highest with the lowest decile and pooling estimates by random-effects meta-analysis. In line with the interpretation as components of total genetic liability, all eight pPRSs were consistently associated with clinical coronary events, with directionally consistent effects across cohorts (**Fig. 3a**, **Supplementary Table 7**). Associations were strongest for the Cardiac remodeling (odds ratio [OR]=1.84, 95% confidence interval [95%CI]:1.56-2.18), Lp(a) (OR=1.59, 95%CI:1.45−1.74) and MUNW (OR=1.56, 95%CI:1.27−1.91) pPRSs, intermediate for the ChEL, TRL, Endothelial dysfunction and Obesity pPRSs (OR range, 1.29−1.42), and modest for the Coagulation pPRS (OR=1.15, 95%CI:1.02−1.29). Each of the eight pPRSs was associated with clinical coronary events, with consistent effect estimates in a population that did not contribute to the GWAS data used for genetic risk decomposition (SCAPIS).

**Figure 3.**
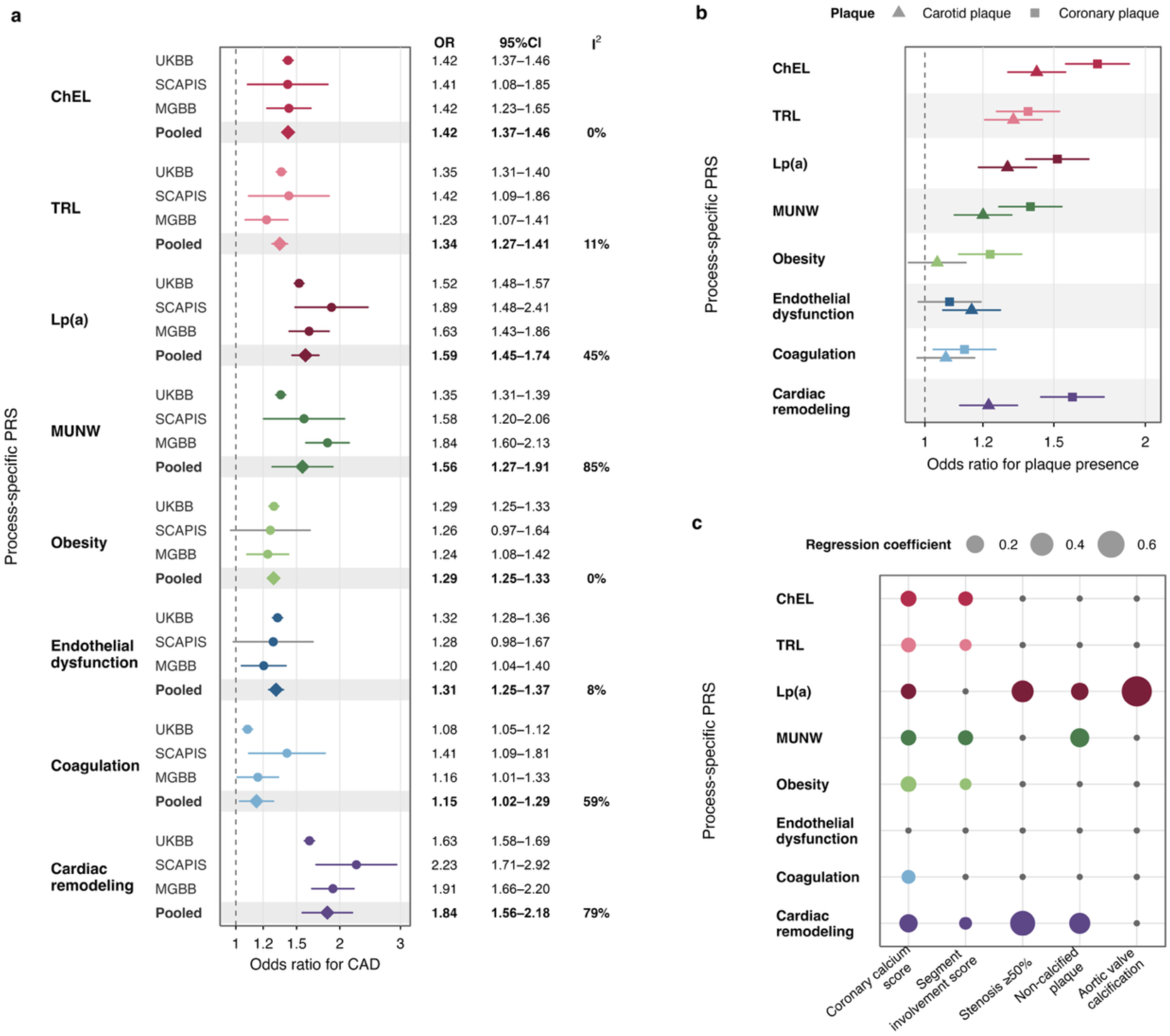
Process-specific polygenic risk scores in relation to subclinical atherosclerosis and clinical coronary events. (**a**) Odds ratios for CAD comparing the highest with the lowest decile of each process-specific polygenic risk score (pPRS) in UKBB, SCAPIS and MGBB, and pooled across cohorts by random-effects meta-analysis. Estimates are from logistic regression adjusted for age, sex and the first ten genetic principal components. (**b**) Odds ratios for the presence of coronary plaque on computed tomography angiography (squares) and carotid plaque on ultrasound (triangles) in SCAPIS, for the highest compared with the lowest decile of each pPRS, adjusted as in Panel a. (**c**) Associations of each pPRS with measures of plaque burden, extent and composition among SCAPIS participants with established coronary plaque. Circle size is proportional to the magnitude of the regression coefficient; colored symbols denote associations reaching nominal statistical significance (P<0.05) and small grey symbols those that did not. Outcomes are coronary calcium score, segment involvement score, obstructive stenosis of 50% or more, non-calcified plaque and aortic valve calcification. Associations with segment involvement score, stenosis and non-calcified plaque were additionally adjusted for coronary calcium score. Abbreviations: ChEL, cholesterol-enriched lipoproteins; Lp(a), lipoprotein(a); MGBB, Mass General Brigham Biobank; MUNW, metabolically unhealthy normal weight; SCAPIS, Swedish CArdioPulmonary bioImage Study; TRL, triglyceride-rich lipoproteins; UKBB, UK Biobank.

We next examined whether these associations differed by sex and ancestry. ChEL, Lp(a), MUNW and Cardiac remodeling pPRSs showed stronger associations with clinical coronary events in men than in women (*P*_Interaction_<0.05; **Extended Data Fig. 3a**, **Supplementary Table 8**). Because the latent genetic components were derived predominantly in participants of European ancestry, we also recomputed each pPRS using the same weights in UKBB participants of self-reported Asian, Black and Mixed/Other ethnicity (**Extended Data Fig. 3b**). In the Asian group (n=1792 cases), associations were largely preserved for seven of the eight pPRSs (OR range, 1.31−1.67), whereas the Obesity pPRS showed no association (OR=1.02, 95%CI:0.84−1.23). In the Black and Mixed/Other groups, estimates were broadly consistent with those in the other groups but less precise, reflecting substantially fewer coronary event cases (n=578 and n=733). Robust characterization in non-European populations will therefore require more diverse studies and ancestry-specific latent genetic component decomposition.

We next tested whether these associations extend to imaging-detected subclinical atherosclerosis in SCAPIS, assessed as coronary plaque on computed tomography angiography and carotid plaque on ultrasound (**Fig. 3b**, **Supplementary Table 9**). Associations were stronger for coronary than for carotid plaque for most pPRSs (OR range 1.13–1.72 versus 1.04–1.42), with the strongest associations for the ChEL (OR=1.72, 95%CI:1.56−1.90), Cardiac remodeling (OR=1.59, 95%CI:1.44−1.76) and Lp(a) pPRSs (OR=1.52, 95%CI:1.37−1.67). These findings suggest that most pPRSs may be more closely related to coronary atherosclerosis than to atherosclerosis in the carotid vascular bed.

Finally, among participants with established coronary plaque, we related each pPRS to measures of plaque burden, extent and composition (**Fig. 3c**, **Supplementary Table 9**). Most pPRSs were associated with coronary artery calcium score and segment involvement score. After adjustment for calcium score, the Cardiac remodeling, Lp(a) and MUNW pPRSs were associated with high-risk features: obstructive stenosis (>50%) with the first two and non-calcified plaque with all. The ChEL pPRS was associated with neither, despite showing the largest association with coronary plaque presence. Aortic valve calcification was associated exclusively with the Lp(a) pPRS. Processes associated with high-risk lesion features were also those most strongly associated with clinical coronary events, suggesting that lesion phenotype might contribute to the differences in risk between processes. These analyses indicate that the eight processes are differently associated with clinical coronary events and subclinical atherosclerosis, and with distinct plaque phenotypes.

### Process-specific polygenic scores capture complex biological processes underlying CAD

We next examined whether the eight pPRSs are associated with distinct cardiometabolic risk profiles, testing each pPRS against a panel of 83 clinical, biochemical and imaging biomarkers in UKBB (**Fig. 4**, **Extended Data Fig. 4**, **Supplementary Table 10**). Each pPRS was associated (P_Bonferroni_<0.05) with a distinct pattern of conventional risk factors (**Fig. 4a**). The lipoprotein-related pPRSs were most strongly associated with their defining lipid fractions (e.g., apolipoprotein B and LDL cholesterol for ChEL, β per s.d. increase in pPRS=0.32 and 0.27). The MUNW and Obesity pPRSs were associated with impaired glycemia (HbA1c, β=0.03 and 0.02), higher liver enzymes (alanine aminotransferase, β=0.04 and 0.02), higher blood pressure (β=0.01–0.02) and subclinical inflammation (C-reactive protein, β=0.01 and 0.02). The Endothelial dysfunction pPRS was associated with impaired kidney function (eGFR_Cystatin_, β=-0.04), higher blood pressure (diastolic, β=0.03) and better glycemic control (HbA1c, β=-0.02). The Coagulation pPRS was associated with glycemia (HbA1c, β=0.03) and higher C-reactive protein levels (β=0.02). The Cardiac remodeling pPRS was associated with better kidney function (eGFR_Cystatin_, β=0.02), higher blood pressure (systolic, β=0.03), and lower C-reactive protein levels (β=- 0.03). Associations of the pPRSs were distinct from the pattern observed for the total CAD polygenic risk score, indicating that they capture distinct risk factor profiles that are diluted in the association pattern of the aggregate score. Replication in SCAPIS and MGBB yielded concordant association patterns (**Supplementary Table 11**).

**Figure 4.**
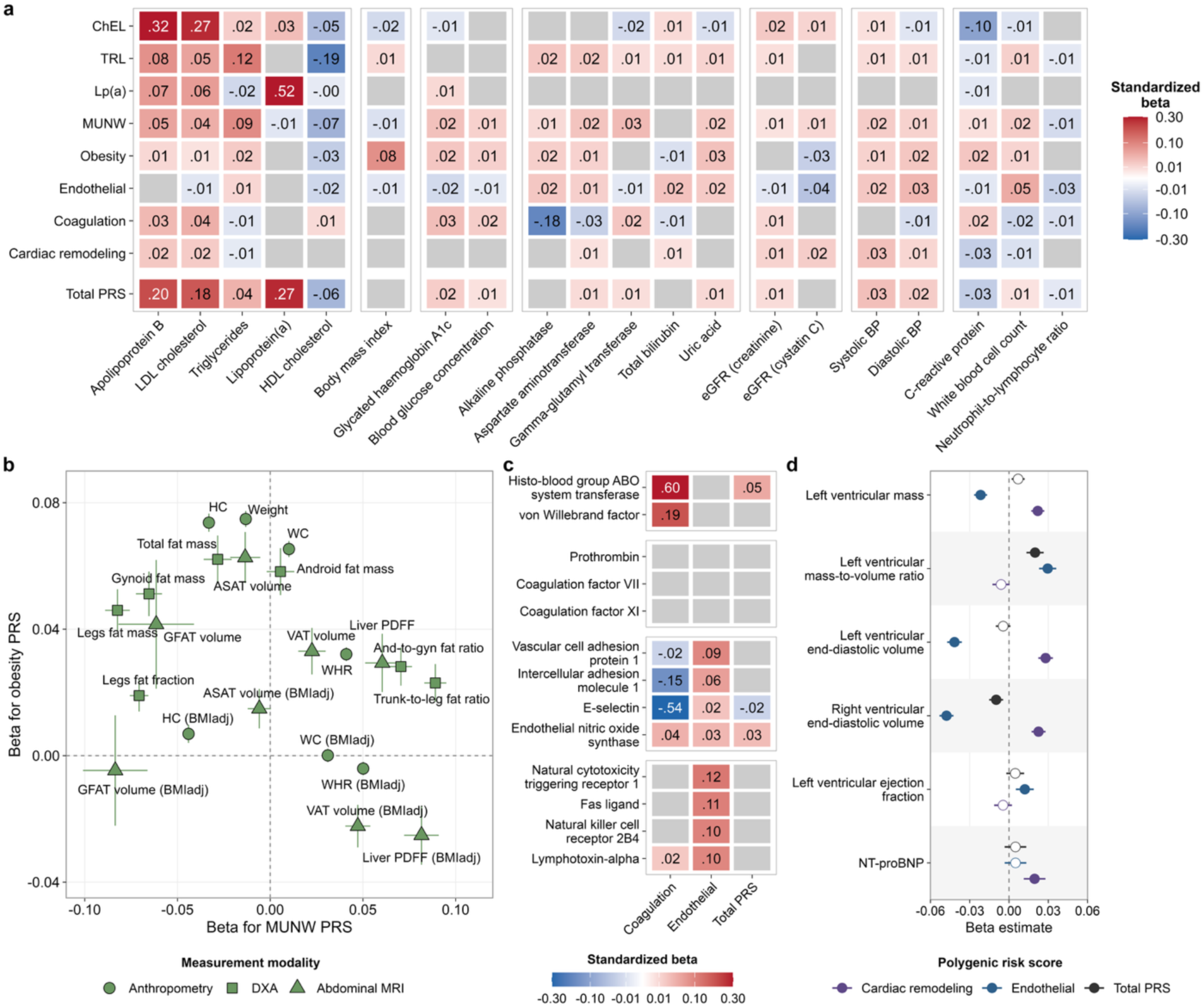
Process-specific polygenic risk scores are associated with distinct biomarker, body composition, proteomic and cardiac profiles. (**a**) Associations of each process-specific polygenic risk scores (pPRS) and of the total CAD PRS with conventional cardiometabolic risk factors in UKBB, grouped by domain: lipids, adiposity and glycemia, liver enzymes, renal function, blood pressure and inflammation. (**b**) Comparison of the MUNW and Obesity pPRS associations across anthropometric, DXA and abdominal MRI measures of body composition. Each point is one measure, positioned by its standardized β for the MUNW pPRS (x-axis) and the Obesity pPRS (y-axis), with whiskers indicating 95% confidence intervals and symbol shape denoting measurement modality. (**c**) Associations of the Coagulation and Endothelial dysfunction pPRSs with selected plasma proteins, grouped by candidate mechanism: the ABO–von Willebrand factor axis, coagulation cascade factors, endothelial activation markers and lymphocyte surface receptors. (**d**) Associations of the Cardiac remodeling pPRS, Endothelial dysfunction pPRS and Total PRS with cardiac magnetic resonance measures of ventricular structure and function, and with NT-proBNP. All estimates are from linear regression adjusted for age, sex and the first ten genetic principal components, with each outcome and each score standardized before analysis. Grey cells in panel a and c indicate associations that did not reach statistical significance after correction for multiple comparisons across all tested markers (P_Bonferroni_<0.05). And-to-Gyn, android-to-gynoid; ASAT, abdominal subcutaneous adipose tissue; BMIadj, adjusted for body mass index; ChEL, cholesterol-enriched lipoproteins; DXA, dual-energy X-ray absorptiometry; eGFR, estimated glomerular filtration rate; GFAT, gluteofemoral adipose tissue; HC, hip circumference; Lp(a), lipoprotein(a); MUNW, metabolically-unhealthy normal weight; PDFF, proton density fat fraction; TRL, triglyceride-rich lipoproteins; UKBB, UK Biobank; VAT, visceral adipose tissue; WC, waist circumference; WHR, waist-to-hip ratio.

Associations with body composition were largely confined to the Obesity and MUNW pPRSs (**Fig. 4b**, **Extended Data Fig. 4**). The Obesity pPRS was broadly associated (P_Bonferroni_<0.05) with measures of body size and with total and compartment-specific adipose tissue content across assessment modalities (β=0.01−0.08). In contrast, the MUNW pPRS was associated with lower total fat mass (β=-0.03) while being positively associated with BMI-adjusted measures of visceral adipose tissue and liver fat (β=0.03−0.08) and inversely with measures of gluteofemoral adipose tissue (β=-0.08−-0.04). These findings support the general adiposity phenotype attributed to the Obesity pPRS and the lipodystrophy-like phenotype attributed to the MUNW pPRS.

We next related each pPRS to 2,923 plasma proteins measured by Olink in UKBB (n = 48,152; **Extended Data Fig. 5**, **Supplementary Table 12**). The proteins most strongly associated with each pPRS were largely consistent with the attributed biology (**Supplementary Table 13**). The Endothelial dysfunction and the Coagulation pPRSs were associated (P_Bonferroni_<0.05) with the largest number of proteins (985 [34%] and 487 [17%], respectively), compared with 32 (1%) for the Cardiac remodeling pPRS. The Coagulation pPRS was associated with ABO and vWF (β=0.60 and 0.19) but not with coagulation cascade proteins (prothrombin, factor VII or XI) (**Fig. 4c**). This pattern suggests that the Coagulation pPRS reflects *ABO*-mediated effects on vWF rather than generalized activation of coagulation. The Endothelial dysfunction pPRS was consistently associated with endothelial activation markers (e.g., vascular cell adhesion protein 1, β=0.09) and, more strongly, with lymphocyte and natural killer cell surface receptors (e.g., natural cytotoxicity triggering receptor 1, β=0.12), implicating this genetic component in impaired endothelial function and vascular inflammation.

Associations with cardiac structure metrics were most pronounced for the Cardiac remodeling pPRS, which was associated (P_Bonferroni_<0.05) with greater left ventricular mass, larger end-diastolic volume and higher stroke volume (all β=0.03) without a change in the mass-to-volume ratio, consistent with eccentric ventricular remodeling (**Fig. 4d**). The Endothelial dysfunction pPRS was associated with the opposite pattern, showing lower left ventricular mass (β=−0.02), smaller end-diastolic volume and lower stroke volume (β=−0.04 for both) and a higher mass-to-volume ratio (β=0.03), consistent with concentric remodeling and a reduced cavity.

Taken together, these analyses indicate that each pPRS carries a distinct biological signature, extending from the traits involved in component extraction to broader patterns of cardiometabolic phenotypes, and that genetic liability to CAD can be resolved into a set of distinguishable biological processes.

### Process-specific polygenic scores are associated with distinct disease profiles

We next investigated whether the eight pPRSs are associated with distinct disease profiles. We tested the association of each pPRS with 30 comorbidities spanning cardiovascular, metabolic, neurological, respiratory, digestive and other diseases, together with cardiovascular death and all-cause mortality, in UKBB (**Supplementary Table 14**). The eight pPRSs showed markedly different comorbidity association patterns (**Fig. 5**).

**Figure 5.**
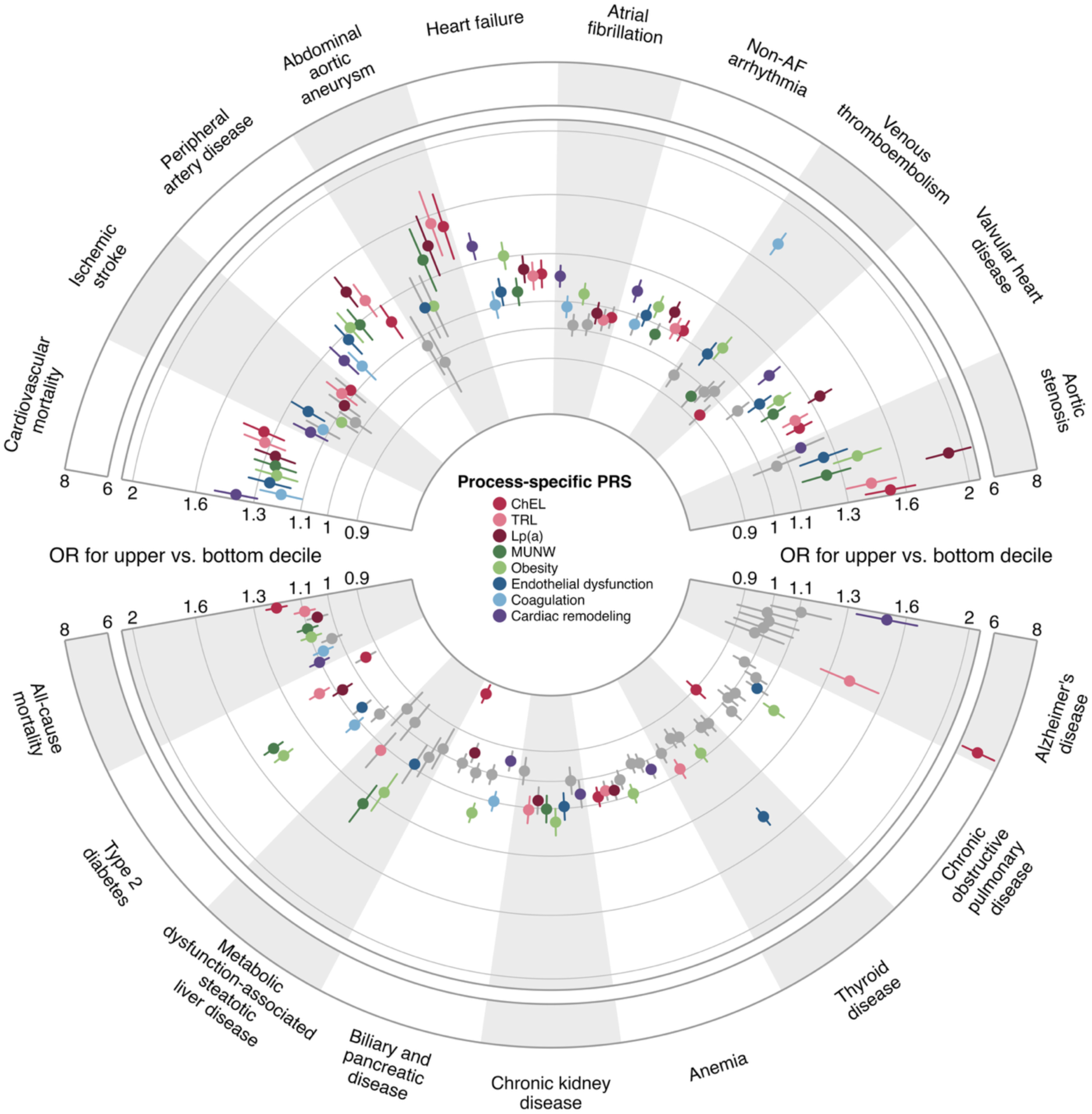
Associations of pPRSs with comorbidities and mortality endpoints. Associations between eight process-specific polygenic risk scores (pPRS) and clinical endpoints in 458,905 UKBB participants. Endpoints included 30 comorbidities spanning cardiovascular, metabolic and neurological disease, together with cardiovascular death and all-cause mortality. Points show odds ratios comparing participants in the top versus bottom decile of each pPRS; horizontal lines show 95% confidence intervals. Results are shown for endpoints associated with at least two pPRSs after Bonferroni correction. Endpoints are grouped into cardiovascular outcomes in the upper half and non-cardiovascular outcomes in the lower half of the diagram. Models were adjusted for age, sex and genetic principal components. ChEL, cholesterol-enriched lipoprotein; Lp(a), lipoprotein(a); MUNW, metabolically unhealthy normal weight; UKBB, UK Biobank.

Most pPRSs were associated (P_Bonferroni_<0.05) with higher risk of non-CAD atherosclerotic outcomes, but the leading process varied by vascular bed. Peripheral artery disease was associated with all eight pPRSs (OR for upper vs. bottom decile range, 1.15−1.45), with the largest estimate for the Lp(a) pPRS (OR=1.45, 95%CI:1.36−1.55). Abdominal aortic aneurysm was associated only with the three lipoprotein-related pPRSs (OR range, 1.45−1.55) and the MUNW pPRS (OR=1.39, 95%CI:1.24−1.57), with the largest estimate for the TRL pPRS (OR=1.55, 95%CI:1.38−1.75). The Endothelial dysfunction pPRS showed the strongest association with ischemic stroke (OR=1.21, 95%CI:1.14−1.30), followed by the Cardiac remodeling pPRS (OR=1.16, 95%CI:1.09−1.24). No pPRS was associated with hemorrhagic stroke.

Among other cardiovascular outcomes, the Cardiac remodeling pPRS was most strongly associated with heart failure and atrial fibrillation (OR=1.38, 95%CI:1.32−1.45; and OR=1.20, 95%CI:1.16−1.25), followed by the Obesity pPRS (OR=1.31, 95%CI:1.25−1.37; and OR=1.14, 95%CI:1.09−1.18). Compared to the other pPRSs, the Coagulation pPRS showed a distinctly larger estimate for venous thromboembolism (OR=1.74, 95%CI:1.67−1.82). The Lp(a) pPRS showed the strongest associations with valvular heart disease (OR=1.35, 95%CI:1.29−1.41) and aortic stenosis (OR=1.90, 95%CI:1.76−2.06).

The associations with metabolic diseases were largely driven by selected pPRSs. T2D and MASLD were associated principally with the body fat-related pPRSs. The largest estimates were for the MUNW pPRS (OR=1.48, 95%CI:1.42−1.54; and OR=1.35, 95%CI:1.25−1.46), followed by the Obesity pPRS (OR=1.46, 95%CI:1.40−1.52; and OR=1.25, 95%CI:1.16−1.35). Chronic kidney disease (CKD) was associated with the Obesity, TRL, MUNW and Endothelial dysfunction pPRSs (OR range, 1.09−1.15).

Three further outcomes were most strongly associated with the ChEL and Obesity pPRSs, in opposing directions for two of them. The ChEL pPRS was inversely associated with biliary and pancreatic disease (OR=0.77; 95%CI:0.75−0.80) and chronic obstructive pulmonary disease (COPD, OR=0.90; 95%CI:0.87−0.94). The Obesity pPRS was positively associated with both (OR=1.16, 95%CI:1.12−1.20; and OR=1.17, 95%CI:1.12−1.22). Both pPRSs were positively associated with anemia (OR=1.07, 95%CI:1.04−1.11; and OR=1.10, 95%CI:1.06−1.13). The Endothelial dysfunction pPRS showed by far the largest estimate for thyroid disease (OR=1.47, 95%CI:1.42−1.53). It was also the only pPRS associated with osteoporosis (OR=1.10, 95%CI:1.05−1.15) and, inversely, with asthma (OR=0.94, 95%CI:0.91−0.96).

Alzheimer’s disease showed a very strong association with the ChEL pPRS (OR=6.93, 95%CI:6.17−7.78), followed by the Cardiac remodeling (OR=1.50, 95%CI:1.34−1.67) and TRL pPRSs (OR=1.41, 95%CI:1.26−1.57). These associations likely reflect in part variants in the *APOE* region, which are established determinants of dementia risk.^33^ In leave-one-out SNP analyses (**Supplementary Table 15**), the association between the ChEL pPRS and Alzheimer’s disease was attenuated after removal of the *APOC1* (−54%) and *APOE* (−9%) variants but persisted. This suggests that the association is not driven solely by these variants. The ChEL pPRS was also associated with Parkinson’s disease (OR=1.20, 95%CI:1.08−1.33).

Cardiovascular death was associated with all eight pPRSs, with similar estimates across processes (OR range, 1.19−1.39). Associations with all-cause mortality were weaker but statistically significant for six pPRSs (OR range, 1.07−1.20).

The total CAD PRS was more strongly associated than any pPRS with atherosclerotic and cardiac outcomes. For most other diseases, individual pPRSs matched or exceeded it, often revealing associations not captured by the total CAD PRS, including inverse associations (**Extended Data Fig. 6**).

Associations with neuropsychiatric and sensory endpoints were modest or absent. The Obesity pPRS was associated with depression (OR=1.06, 95%CI:1.03−1.10) and the Cardiac remodeling pPRS with anxiety disorders (OR=1.07, 95%CI:1.03−1.11). No pPRS was associated with schizophrenia, migraine or headache, epilepsy, hearing loss or glaucoma.

### Risk load in specific pPRSs modifies the contribution of dynamic factors to CAD risk

Finally, we examined whether the pPRSs identify individuals in whom conventional risk factors carry greater risk due to their genetic liability. We tested interactions between each pPRS and six conventional risk factors (LDL cholesterol, remnant cholesterol, systolic blood pressure, HbA1c, high-sensitivity C-reactive protein, and waist circumference) for incident myocardial infarction among participants free of atherosclerotic cardiovascular disease at baseline (**Extended Data Fig. 7**, **Supplementary Table 16**).

Interaction terms were significant (P_Bonferroni_<0.05) for three pPRS-risk factor pairs: the MUNW pPRS with waist circumference, the ChEL pPRS with LDL cholesterol, and the TRL pPRS with remnant cholesterol. After further comprehensive covariate adjustment, the ChEL and TRL interactions were rendered statistically non-significant (P_Interaction_=0.053 and P_Interaction_=0.10), whereas the interaction between the MUNW pPRS and waist circumference remained statistically significant (P_Interaction_=5×10^-4^). The association between waist circumference and incident myocardial infarction was stronger at higher levels of the MUNW pPRS (**Fig. 6a**). The equivalent interaction was not significant for the total CAD polygenic risk score (P_Interaction_=0.46) (**Fig. 6b**).

**Figure 6.**
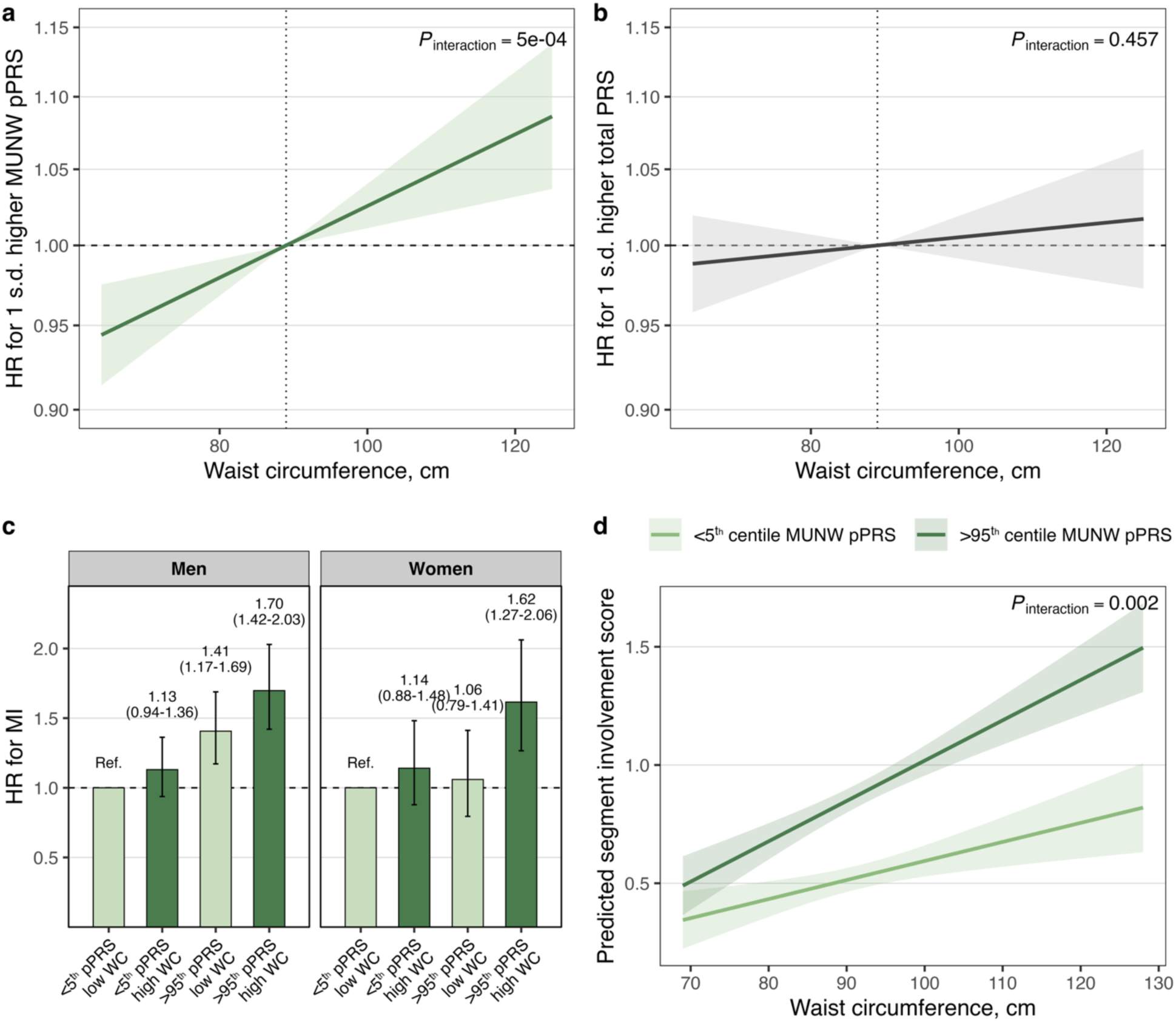
The MUNW process-specific polygenic risk score modifies the association between abdominal adiposity and incident myocardial infarction. (**a**) Hazard ratio for myocardial infarction per 1-s.d. higher MUNW process-specific polygenic-risk-score (pPRS) across the range of waist circumference, estimated from a Cox model including a pPRS × waist circumference interaction term in UKBB. The solid line shows the estimated hazard ratio and the shaded band its 95% confidence interval. P_Interaction_ reflects likelihood ratio test of the interaction term. (**b**) The corresponding analysis for the total CAD polygenic risk score, modelled and displayed as in a. (**c**) Hazard ratios for myocardial infarction across joint categories of MUNW pPRS and waist circumference, shown separately for men and women in UKBB. Participants with both a low pPRS and a low waist circumference serve as the reference category. (**d**) Predicted segment involvement score on coronary computed tomography angiography across the range of waist circumference, for participants below the 5th and above the 95th centile of the MUNW pPRS in SCAPIS. Lines show predictions from a linear model including a pPRS × waist circumference interaction term with 95% confidence bands. MUNW, metabolically unhealthy normal weight; SCAPIS, Swedish CArdioPulmonary bioImage Study; UKBB, UK Biobank.

In sex-stratified analyses, the joint categories of MUNW pPRS and waist circumference showed a gradient in both men and women, with the highest risk among those in the high categories of both (hazard ratio [HR]=1.70, 95%CI 1.42–2.03 in men; HR=1.62, 95%CI:1.27–2.06 in women) relative to participants low on both (**Fig. 6c**). Neither a high pPRS nor a high waist circumference alone conferred comparable risk. In SCAPIS, we then tested an interaction between MUNW pPRS and waist circumference on subclinical atherosclerosis (**Fig. 6d**). The association between waist circumference and segment involvement score was stronger in participants in the top category than in the lowest category of the MUNW pPRS (P_Interaction_=0.002).

Our results indicate that genetic susceptibility to adverse adipose tissue distribution shapes the association between abdominal adiposity and incident myocardial infarction. These findings identify MUNW genetic liability as a candidate modifier of the relationship between abdominal adiposity and CAD risk, providing testable hypotheses for intervention studies.

## Discussion

In this study, we conducted data-driven latent component decomposition of CAD genetic risk variants based on their profile of association with 78 CAD-relevant phenotypic traits. We identified eight genetic components, delineating potentially separable biological axes underlying CAD risk, related to cholesterol-enriched lipoproteins, triglyceride-rich lipoproteins, lipoprotein(a), metabolically unhealthy normal weight, obesity, endothelial dysfunction, coagulation and cardiac remodeling. Chromatin accessibility enrichment mapped the components to cell types consistent with their biological process interpretation. We derived pPRSs reflecting an individual’s risk load on each genetic axis. These scores were associated with distinct profiles of clinical, molecular and imaging biomarkers, and with distinct patterns of disease endpoints. We also provided evidence that the CAD risk axis related to adverse adipose tissue distribution modifies the association between abdominal obesity and incident myocardial infarction.

Retention of apoB-containing lipoproteins in vascular walls is the primary cause of atherosclerosis.^17,19^ Among the eight data-driven latent genetic CAD risk components, three components reflect biological determinants of LDL, TRL, and Lp(a), major apoB-containing lipoprotein classes. This separation is in line with increasing evidence that TRL and Lp(a) contribute to atherosclerosis independently of LDL.^34–36^ The pPRSs for ChEL, TRL and Lp(a) included variants at loci governing the metabolism of the respective lipoprotein classes, and each pPRS was associated most strongly with circulating concentrations of the corresponding lipoprotein. These axes therefore separate genetic liability for each lipoprotein class. While existing lipid-lowering therapies such as statins and PCSK9 inhibitors primarily lower LDL, emerging therapies target TRL and Lp(a). The quantification of individual risk load in these genetic axes may provide clinically useful information beyond directly measured lipoprotein concentrations, because inherited liability across several lipoprotein-related processes appears to be related to distinct co-morbidity patterns and suggests treatment-response hypotheses that can be tested in intervention studies that target specific mechanisms in lipoprotein metabolism.

We identified two genetic components reflecting CAD risk related to distinct adipose tissue phenotypes. The Obesity component reflected general adiposity and body size, whereas MUNW reflected a lipodystrophy-like pattern of fat redistribution, with lower total and gluteofemoral fat but greater visceral and hepatic fat content. These components are in line with prior genetic studies showing that adiposity-increasing variants separate into groups promoting general fat accumulation and groups promoting reduced lower-body fat with ectopic deposition in the liver.^16,21,37^ In those studies the direction of fat storage determined the metabolic consequences of adiposity, with variants favoring subcutaneous storage conferring lower risk of T2D, hypertension and CAD despite higher body fat. Similarly, genetic latent factor models in T2D have separated genetic components for general obesity and for lipodystrophy-like fat distribution.^11–14^ In our study both the MUNW and Obesity pPRSs were associated with adverse cardiometabolic biomarkers and with higher risk of all-cause mortality, T2D, MASLD and CKD. However, atrial fibrillation, venous thromboembolism, COPD and biliary disease were associated with the Obesity pPRS but not with MUNW. Adiposity therefore contributes to CAD risk through two distinct biological processes that partially converge on metabolic consequences and are related to different disease risk patterns.

The Endothelial dysfunction component captured variants at loci related to endothelial function and vascular inflammation. This interpretation was supported by nominal enrichment in endothelial cell types and by proteomic associations with markers of endothelial activation and with receptors of lymphocyte and natural killer cell activation. Notably, despite its association with clinical CAD, this was the only component not associated with coronary plaque presence, which may indicate that it confers risk through vascular mechanisms other than atherosclerotic plaque burden. In a hypothesis-based approach, a recent study constructed a curated endothelial-cell PRS that predicted CAD independently of the total CAD PRS.^38^ Our latent genetic component decomposition identified a comparable axis through a data-driven approach.

We identified a separate genetic component mapping to thrombosis-related CAD risk, dominated by the *ABO* locus. Its pPRS was associated with ABO glycosyltransferase, vWF and glycan-binding proteins, and inversely with soluble endothelial adhesion molecules, but not with coagulation cascade factors. This pattern is consistent with prior evidence that *ABO*-dependent glycosylation determines the circulating concentrations of vWF and related glycoproteins and thereby contributes to thrombotic risk.^39,40^ An analogous component defined by the same locus has been identified in genetic component decomposition of T2D.^12,13^ Although driven largely by one locus, the elevated risk of both, clinical coronary events and venous thromboembolism, related to high risk load in this component is of potential clinical relevance; notably, the total CAD PRS was not associated with venous thromboembolism.

Finally, a Cardiac remodeling genetic component emerged with clear linkage to cardiac structure-related phenotypes and pronounced CAD risk associations. Its variants were most strongly enriched in regulatory elements of arterial vascular smooth muscle cells, and further in endocardial cells, pericytes and cardiac fibroblasts, but not in cardiomyocytes, implicating the non-myocyte compartment in which fibrotic remodeling originates. In downstream analyses the Cardiac remodeling pPRS was associated with left ventricular structure in a pattern indicating eccentric remodeling. The Cardiac remodeling component showed the strongest association with clinical CAD of any pPRSs and among the strongest associations with subclinical atherosclerosis. In line with its relation to myocardial structure, it also showed strong associations with heart failure and atrial fibrillation. Previous studies have shown that left ventricular structure predicts cardiovascular events independently of conventional risk factors.^41^ Impaired coronary microvascular function accompanies myocardial remodeling and may itself raise the risk of ischemic cardiac events.^42^ The extraction of this less expected genetic risk component underpins the advantage of our unbiased data-driven approach; mechanistic validation studies are warranted.

We identified a significant interaction between the MUNW pPRS and waist circumference, indicating that individuals with high risk load in this genetic axis may be more vulnerable to the harmful effects of abdominal adiposity. Notably, this interaction was replicated for subclinical atherosclerosis associations in an independent study population, but no interaction was observed between waist circumference and the total CAD polygenic score. Our findings are in line with the adipose expandability hypothesis, in which lipids are deposited ectopically once subcutaneous storage capacity is exceeded.^43^ In T2D, lipodystrophy-related pPRS accounted for much of the difference in risk-equivalent BMI between ancestry groups, indicating that genetically-informed fat distribution can refine adiposity thresholds.^13^ These findings provide a potential example for the application of pPRSs in precision prevention of CAD: visceral adiposity reduction may be particularly important for individuals with high risk load in the MUNW pPRS.

Our study has several strengths. We conducted data-driven latent component decomposition of genetic CAD risk variants across a broad panel of relevant phenotypes, without specifying the processes in advance. Chromatin accessibility data that were not used in the component decomposition corroborated the biological interpretation of the resulting genetic risk axes. We also tested the derived pPRSs against clinical CAD in three large biobanks, and characterized their associations across biochemical, proteomic and imaging phenotypes in more than 500,000 deeply phenotyped participants.

Several limitations apply to our study. Our data-driven approach relies on available data, and the detection and biological interpretation of the components partially reflect the strength of phenotype and cell type data as well as genes that are linked to each biological axis underlying the components. The applied latent component decomposition approach only considered variants reaching genome-wide significance of CAD risk association; a larger set of variants might resolve additional mechanisms. The genetic components were derived in participants of predominantly European ancestry. Although seven of eight pPRSs were reproduced in participants of Asian ethnicity, ancestry-specific trait matrix curation and factor decomposition may be required for other groups. Finally, the interactions between modifiable risk factors and the pPRSs were observational, and confirmation will require repeated measurement of risk factors over time or evidence from intervention trials.

We show that genetic liability to CAD can be decomposed into eight biologically distinct components that capture different contributions to an individual’s overall genetic risk. These process-specific polygenic risk scores can distinguish individuals with similar overall genetic liability according to the biological processes contributing to that risk. Their divergent associations with phenotypic traits, circulating biomarkers, subclinical disease characteristics and comorbidities demonstrate that similar levels of genetic CAD risk can arise through different biological processes. The interaction between the ectopic fat deposition-related component and visceral adiposity further provides a potential route toward biologically targeted prevention by identifying individuals in whom fat distribution-related pathways may be particularly relevant. This framework moves polygenic risk assessment beyond quantification of overall liability toward a biologically resolved representation of genetic CAD risk that can be tested for precision prevention and management.

## Methods

### Latent decomposition of CAD risk variants

We extracted variants associated with CAD at genome-wide significance (P < 5×10^-8^) from seven GWAS of predominantly European ancestry.^6,44–48^ A variant was retained if it reached this threshold in at least one contributing study, and its effect estimate and standard error were taken from the study with the largest sample size in which it was reported. We excluded insertions and deletions and variants in the major histocompatibility complex, whose extended haplotype structure precludes reliable pruning. The remaining variants were thinned to an approximately independent set by linkage disequilibrium (LD) pruning (r^2^ < 0.05; minor allele frequency > 0.001) against the European reference panel.

We assembled summary statistics for 87 continuous cardiometabolic phenotypes and allowed the pipeline to determine which carried information about the CAD variant set rather than specifying biological processes in advance. For each trait, we selected the GWAS with the largest available sample size, preferring studies of European-ancestry participants and using multi-ancestry studies where no European-ancestry GWAS was available. Trait GWAS with a median sample size below 5,000 or excessive missingness across the pruned CAD variants were excluded.

We next assessed variant coverage across the trait GWAS. Variants with association statistics available for fewer than 80% of traits, as well as multiallelic and strand-ambiguous variants (A/T or C/G), were replaced where possible by proxies in strong LD in the European reference population (r^2^ > 0.8) that had adequate trait coverage. Variants for which no suitable proxy could be identified were excluded. Each retained variant was oriented to its CAD risk-increasing allele, yielding 526 variants for decomposition.

For each variant–trait association, we calculated a Z score by dividing the effect estimate by its standard error. Residual missing Z scores were imputed using low-rank matrix completion implemented in *softImpute* package,^49^ with the regularization parameter selected by masking 10% of observed entries and minimizing the root-mean-square imputation error. Traits were subsequently excluded if no variant reached Bonferroni significance (P < 0.05/526) or if they were highly correlated with another trait (Pearson |r| > 0.80), retaining the trait with the stronger maximum variant association. This left 78 traits. Associations were signed relative to the CAD risk-increasing allele. Because non-negative factorization cannot represent signed inputs, each trait was represented by separate positive and negative columns, with the unused direction set to zero, giving a final input matrix of 526 variants × 156 signed trait columns.

We then identified components of variants sharing a common trait signature using bNMF, which decomposes the input into two matrices: one of trait weights and one of variant weights, each with one column per component. The method performs soft clustering: rather than assigning each variant to a single component, it gives every variant a weight on every component, so that loci with pleiotropic effects contribute to more than one process. Previous applications of this framework have used a single weight threshold across all components to select top-weighted variants. We refined this method by determining a component-specific threshold at the inflection point of each component’s ranked weight distribution, identified using the unit invariant knee method. The algorithm was run from 50 random initializations, and the number of components was taken as the rank recovered most frequently across runs.

### Chromatin accessibility enrichment analysis

For each component we defined 99% credible sets around its constituent index variants. Within a 500-kb window of each index variant, all variants in linkage disequilibrium at r^2^ ≥ 0.1 were assigned an approximate Bayes factor using the Wakefield method,^50^ with a prior standard deviation of 0.2 on the log odds ratio. Posterior probabilities of association were obtained by dividing each variant’s Bayes factor by the sum of Bayes factors at that locus, and the credible set was defined as the smallest set of variants whose cumulative posterior probability reached 0.99.

Chromatin accessibility was assessed using single-nucleus ATAC-seq data from three atlases: 1,143,424 candidate cis-regulatory elements across 111 adult cell types,^51^ a dedicated human heart atlas of 287,141 elements across nine cardiac cell types,^52^ and a coronary artery atlas of eight cell types.^53^

Enrichment was tested with g-chromVAR (*gchromVAR* package),^54^ which scores each cell type by the overlap of credible-set variants, weighted by their posterior probabilities, against a background matched on guanine-cytosine content and total accessibility. One-sided enrichment P values were obtained from the upper tail of the z-scores. To account for multiple testing, we applied a Bonferroni-corrected threshold of P < 3.9×10^-4^ (0.05 / 128 cell types).

### Derivation of process-specific polygenic risk scores

For each component we derived a pPRS as the sum of CAD risk-allele dosages, each weighted by the variant’s bNMF weight for that component. Scores were computed from imputed genotype dosages and standardized to mean zero and unit variance within each cohort. For comparison we constructed a total CAD polygenic risk score as the unweighted sum of risk-allele dosages across 196 variants contributing to at least one bNMF component.

### Study sources

#### UK Biobank

UKBB is a prospective cohort of approximately 500,000 adults aged 40 to 69 years at enrolment, recruited between 2006 and 2010 at 22 assessment centers across the United Kingdom.^55^ Participants completed baseline questionnaires and underwent physical measurement and blood sampling, with a subset returning from 2014 onwards for multimodal imaging assessment.^56^ Genotyping was performed on the UK Biobank Axiom and UK BiLEVE arrays with imputation to the Haplotype Reference Consortium and UK10K reference panels. Analyses were restricted to 458,905 participants of self-reported White ancestry. Baseline characteristics of these participants are presented in the **Supplementary Table 17**.

#### Swedish CArdioPulmonary bioImage Study

SCAPIS is a population-based cohort of 30,154 adults aged 50 to 64 years, randomly recruited from six Swedish university hospital catchment areas between 2013 and 2018.^57^ At the baseline visit participants underwent coronary computed tomography angiography and carotid ultrasound, together with clinical and biochemical characterization. Genotyping was performed on a customized Illumina GSA-MDv3 array, with imputation to the Haplotype Reference Consortium r1.1 panel. We restricted our analysis to 24,904 participants of European-ancestry, who had available genotyping data (24,835 participants had registry-linkage data for outcome analysis). Baseline characteristics of these participants are presented in the **Supplementary Table 18**.

#### Mass General Brigham Biobank

MGBB is a health-system-based biobank of consented patients across the Mass General Brigham health system, with genotype data linked to longitudinal electronic health records.^58^ Genotyping was performed on the Illumina Global Screening Array in four batches, with imputation to the TOPMed r2 multi-ancestry reference panel at the TOPMed Imputation Server using Eagle and Minimac4. Analyses were restricted to participants of genetically inferred European ancestry who met study-specific quality-control and phenotype-availability criteria (n = 53,848). Baseline characteristics of these participants are presented in the **Supplementary Table 19**.

### Associations of process-specific scores with clinical coronary events

Associations between the pPRSs and CAD were analyzed in a case-control setting. In UK Biobank, clinical coronary events were defined as (1) self-reported myocardial infarction or coronary revascularization, (2) hospital inpatient diagnosis of or death from coronary artery disease, or (3) hospitalization for coronary artery bypass grafting or percutaneous coronary intervention (provided with the analysis code; see Code Availability). In SCAPIS, clinical coronary events were defined as (1) self-reported myocardial infarction in the baseline questionnaire or (2) hospital inpatient diagnosis of or death from myocardial infarction (see Code Availability). In the Mass General Brigham Biobank, clinical coronary events were defined using a previously validated phenotyping algorithm (“CAD” phenotype) applied to electronic health records,^58^ with a separate definition applied to confirm non-cases.

In each cohort, each pPRS was tested against clinical CAD in logistic regression models adjusted for age, sex and the first ten genetic principal components. Estimates were rescaled by a factor of 3.51 to express the contrast between the highest and lowest deciles of each score. Cohort-specific estimates were pooled by random-effects meta-analysis with restricted maximum likelihood estimation of the between-study variance. I^2^ statistic was calculated to quantify proportion of total variability due to between-study heterogeneity.

### Associations of process-specific scores with subclinical atherosclerosis measures

We analyzed cross-sectional associations between the pPRSs and measures of subclinical atherosclerosis in SCAPIS. Participants underwent coronary computed tomography angiography under a standardized protocol on a dual-source scanner (Somatom Definition Flash, Siemens Medical Solutions), read using an 18-segment coronary model, as described previously.^57^ We included six derived measures: coronary plaque presence (binary; plaque in at least one segment), coronary artery calcium score (Agatston score summed across segments), segment involvement score (number of segments containing plaque), obstructive stenosis (any segment with 50% or greater stenosis), non-calcified plaque presence (binary; non-calcified plaque in any segment) and aortic valve calcification. Carotid plaque presence was defined as plaque in the left or right carotid artery on baseline ultrasound (Acuson S2000, Siemens).^57^

Associations between the pPRSs and binary outcomes were analyzed by logistic regression and continuous outcomes by linear regression. The coronary artery calcium score was log(x+1) transformed to correct for right skewness. All models were adjusted for age, sex and the first ten genetic principal components; models for the segment involvement score, non-calcified plaque and obstructive stenosis were additionally adjusted for the coronary artery calcium score.

### Associations of process-specific scores with clinical, biochemical and imaging biomarkers

We analyzed associations between the pPRSs, the total CAD PRS and a curated set of 83 clinical, biochemical and imaging biomarkers in UKBB. Clinical measurements and biochemistry were obtained at the baseline assessment visit, whereas body composition by dual-energy X-ray absorptiometry, abdominal magnetic resonance imaging and cardiac magnetic resonance imaging were acquired at a separate imaging visit, as described previously.^56^ Details of the specific variables and field codes used are provided with the analysis code (see Code availability). Associations were analyzed in linear regression models adjusted for age, sex and the first ten genetic principal components. To account for multiple testing, we applied a Bonferroni-corrected threshold of P< 6.0×10^-4^ (0.05 / 83 biomarkers). Of these biomarkers, 39 were also available in SCAPIS and/or MGBB for replication of pPRS–biomarker associations and were modeled using the same approach. Associations were pooled across cohorts by random-effects meta-analysis with restricted maximum likelihood estimation.

We analyzed associations of the pPRSs with circulating plasma proteins in UKBB. Plasma proteomic data were generated using the Olink Explore 3072 platform, which employs proximity extension assay technology to quantify the relative abundance of 2923 proteins. Associations between each score and protein abundance were analyzed in linear regression models adjusted for age, sex and the first ten genetic principal components. To account for multiple testing, we applied a Bonferroni-corrected threshold of P<1.7×10^-5^ (0.05 / 2923 proteins).

### Associations of process-specific scores with comorbidities and mortality

We analyzed associations of the pPRSs and the total CAD PRS with 30 comorbidities, together with cardiovascular death and all-cause mortality, in UKBB. Outcomes were analyzed in a case-control setting, with definitions provided with the analysis code (see Code availability). Associations were analyzed in logistic regression models adjusted for age, sex and the first ten genetic principal components. To account for multiple testing, we applied a Bonferroni-corrected threshold of P<1.6×10^-3^ (0.05 / 32 outcomes).

### Interactions of process-specific scores with conventional cardiovascular risk factors

We tested multiplicative interactions between each pPRS and six conventional risk factors (LDL cholesterol, remnant cholesterol, systolic blood pressure, HbA1c, high-sensitivity C-reactive protein, and waist circumference) in relation to incident myocardial infarction in UKBB. The definition of incident myocardial infarction is provided with the analysis code (see Code availability). Among participants free of atherosclerotic cardiovascular disease at baseline, we fitted Cox proportional hazards models containing each pPRS, the risk factor and their product term, adjusted for age, sex and the first ten genetic principal components. Interactions were tested by likelihood ratio test and Bonferroni-corrected P<0.008 (0.05/6 risk factors). Interaction pairs reaching Bonferroni-corrected significance were re-tested in comprehensively adjusted models additionally including body mass index, smoking, remaining risk factors and estimated glomerular filtration rate. Because the interaction between the MUNW pPRS and waist circumference was the only one to remain significant, we estimated hazard ratios for myocardial infarction across joint categories of waist circumference and MUNW pPRS, separately in men and women. As external validation, we tested the interaction between the MUNW pPRS and waist circumference in relation to the segment involvement score in SCAPIS, modeled continuously, in linear regression adjusted for age, sex, the first ten genetic principal components and body mass index.

## Data Availability

This study used data from the UK Biobank (UKBB), the Swedish CArdioPulmonary bioImage Study (SCAPIS) and the Mass General Brigham Biobank (MGBB). These datasets contain sensitive participant-level data and are governed by approved data access policies that meet ethical and data security requirements, so the authors cannot share them publicly. UKBB data are available to bona fide researchers through a formal application process via the UKBB Access Management System (https://www.ukbiobank.ac.uk), and this research was conducted under UKBB application numbers 570811. SCAPIS data are available to researchers based in Sweden, or to international researchers connected to a Swedish research institution, through applications submitted via the SCAPIS data portal (https://www.scapis.org); applications are assessed case by case and require ethical approval from the Swedish Ethical Review Authority. MGBB data are available on request to the Mass General Brigham Biobank (https://research.massgeneralbrigham.org/en/researchers-and-collaborators/personalized-medicine/biobank), subject to institutional review board approval. All GWAS summary statistics used for the genetic decomposition are publicly available and are cited in Supplementary Tables 1 and 2. Non-sensitive, summary-level data underlying the study results are provided in the manuscript and the accompanying code repository (see Code availability).

## Declarations

### Ethics approval

The genetic decomposition used only publicly available summary-level data from previously published genome-wide association studies, each of which obtained ethical approval and participant consent. The North West Multi-Centre Research Ethics Committee of the NHS Health Research Authority gave ethical approval for the UK Biobank (reference 11/NW/0382). The Regional Ethical Review Board in Umeå, Sweden, gave ethical approval for the Swedish CArdioPulmonary bioImage Study (SCAPIS) (Dnr 2010-228-31M). The Swedish Ethical Review Authority gave ethical approval for this work (Dnr 2023-07277-01). The Institutional Review Board of Mass General Brigham, Boston, MA, USA, gave ethical approval for this work (protocol 2016P001018). All participants in the UK Biobank, SCAPIS and the Mass General Brigham Biobank provided written informed consent.

### Data availability

This study used data from the UK Biobank (UKBB), the Swedish CArdioPulmonary bioImage Study (SCAPIS) and the Mass General Brigham Biobank (MGBB). These datasets contain sensitive participant-level data. Approved data access policies govern them and meet ethical and data security requirements, so the authors cannot share them publicly. UKBB data are available to bona fide researchers through a formal application via the UKBB Access Management System (https://www.ukbiobank.ac.uk). This research was conducted under UKBB application number 570811. SCAPIS data are available to researchers based in Sweden, or to international researchers connected to a Swedish research institution, through applications submitted via the SCAPIS data portal (https://www.scapis.org). Applications are assessed case by case and require ethical approval from the Swedish Ethical Review Authority. MGBB data are available on request from the Mass General Brigham Biobank (https://research.massgeneralbrigham.org/en/researchers-and-collaborators/personalized-medicine/biobank), subject to institutional review board approval. All GWAS summary statistics used for the genetic decomposition are publicly available and are cited in Supplementary Tables 1 and 2. Non-sensitive, summary-level data underlying the study results are provided in the manuscript, the Supplementary Information and the accompanying code repository (see Code availability).

### Code availability

The bNMF pipeline and the analysis code, including phenotype definitions, are available at [TBA].

---

**Extended Data Figure 1.**
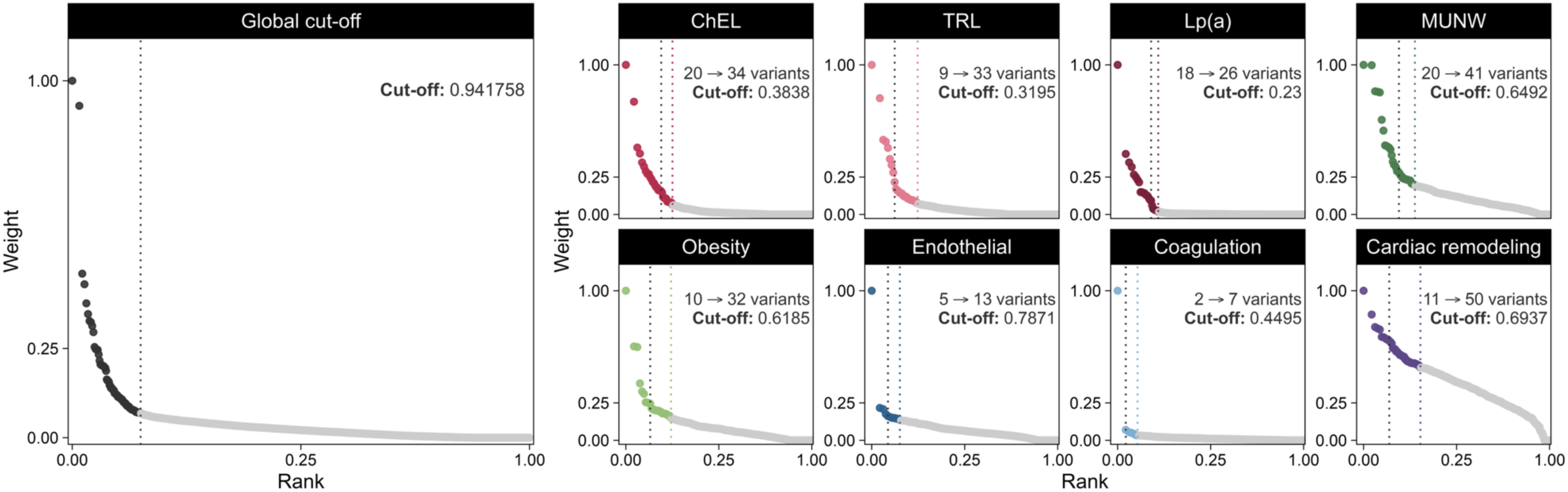
Comparison of variants retained per component using global and component-specific thresholds. Variant weight is plotted against rank (both scaled 0–1) for each bNMF component; the elbow of the ranked-weight curve defines the inclusion threshold for top-weighted variants. (Left) A single global threshold (weight = 0.94) applied uniformly across all components, shown on the pooled ranked-weight distribution. (Right) Component-specific thresholds selected independently for each of the eight CAD components by the unit-invariant knee (UIK) method, which identifies the point of maximum curvature of each component’s ranked-weight curve. ChEL, cholesterol-enriched lipoproteins; Lp(a), lipoprotein(a); MUNW, metabolically unhealthy normal weight; TRL, triglyceride-rich lipoproteins.

**Extended Data Figure 2.**
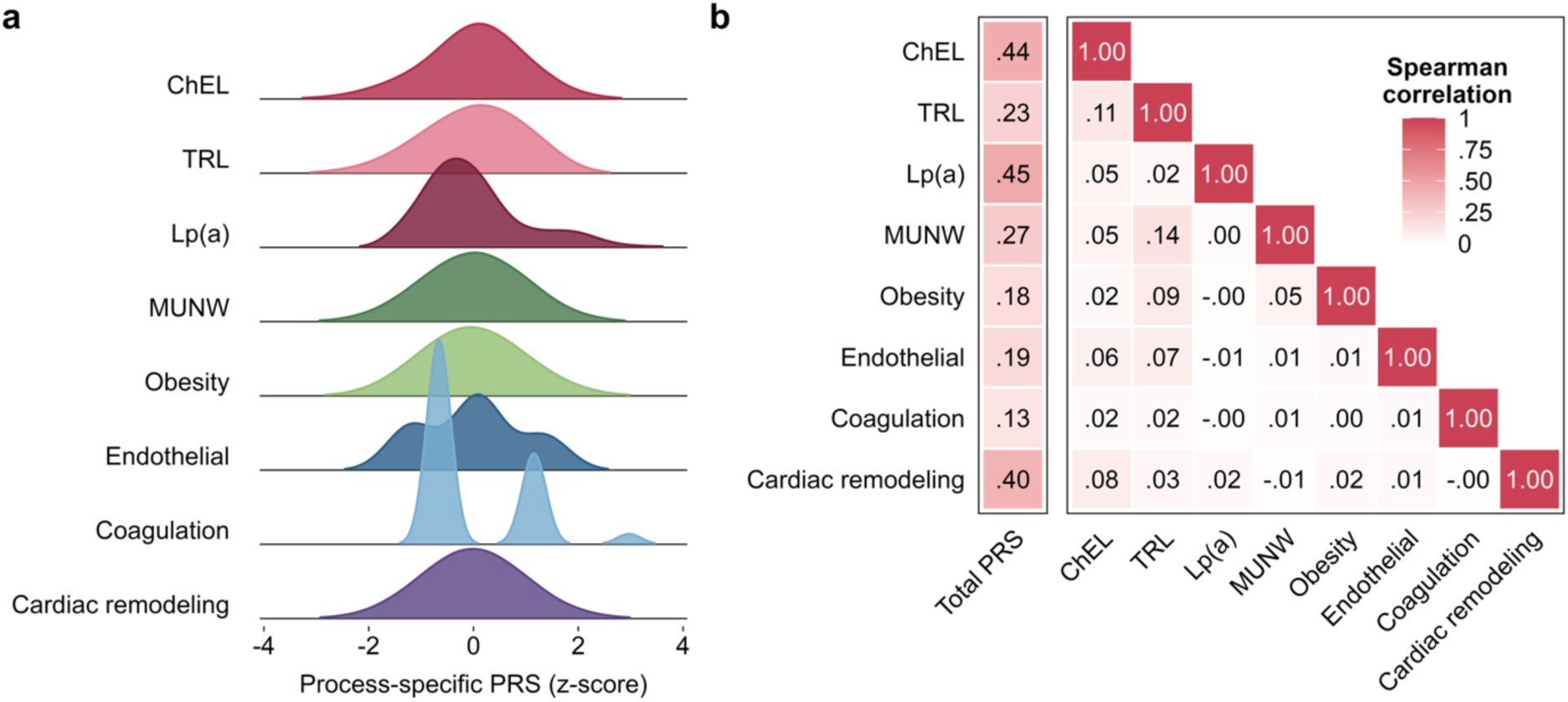
Distribution and correlations of the component process-specific polygenic risk scores. (**a**) Distribution of each component process-specific polygenic risk scores (pPRS) across UKBB participants of White ancestry (n = 458,905) shown as z-score-standardized densities (ridgeline plot). (**b**) Spearman rank correlations among the scores. The left column shows the correlation of each component pPRS with the total PRS for CAD; the right matrix shows pairwise intercorrelations between the eight component pPRS. Color intensity encodes the correlation coefficient (scale at right). ChEL, cholesterol-enriched lipoproteins; Lp(a), lipoprotein(a); MUNW, metabolically unhealthy normal weight; TRL, triglyceride-rich lipoproteins; UKBB, UK Biobank.

**Extended Data Figure 3.**
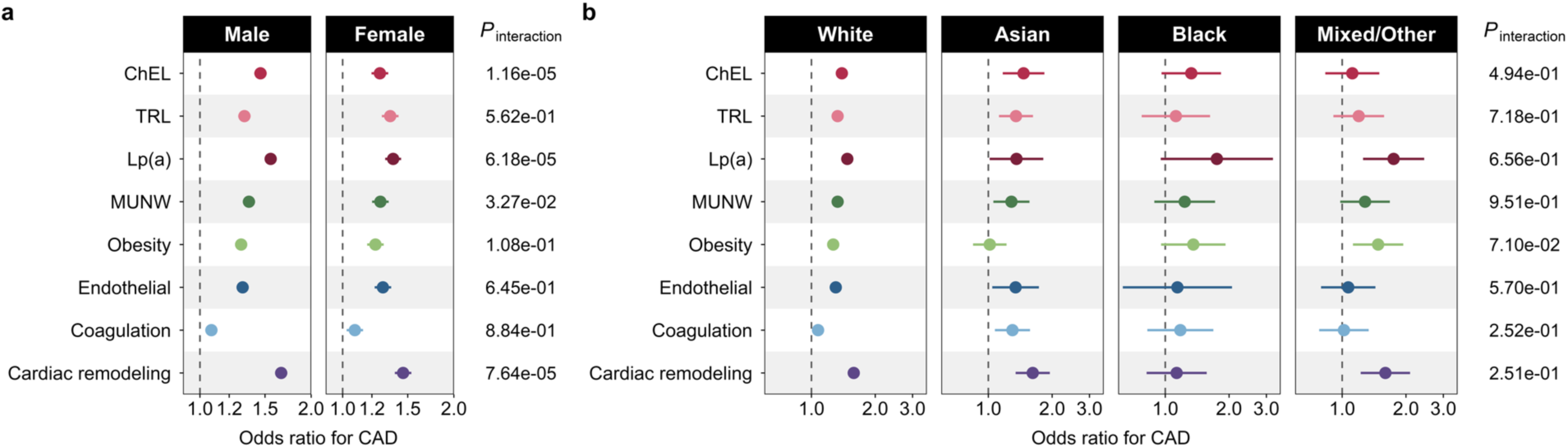
Sex- and ethnicity-stratified associations of the process-specific polygenic risk scores with clinical coronary events. Associations between each process-specific polygenic score (pPRS) and clinical coronary events in UKBB. Points are odds ratios comparing the top versus the bottom decile of each pPRS, with horizontal bars showing 95% confidence intervals. All estimates and interaction P values were derived from interaction models, adjusted for age, sex, and the first ten genetic principal components. **(a)** Sex-specific estimates, modeled with a pPRS × sex interaction term and female participants as the reference group. **(b)** Ethnicity-specific estimates, modeled with a pPRS × ethnicity interaction term and White participants as the reference group. For panel (b), we calculated pPRS in participants of Asian (n = 10,908), Black (n = 7631), and Mixed/Other (n = 7387) ethnicity using the component weights derived from the main analysis. In both panels, *P*_interaction_ indicates likelihood-ratio test comparing the full model, including the pPRS × stratum interaction term, against the reduced model without it. ChEL, cholesterol-enriched lipoproteins; TRL, triglyceride-rich lipoproteins; Lp(a), lipoprotein(a); MUNW, metabolically unhealthy normal weight; UKBB, UK Biobank.

**Extended Data Figure 4.**
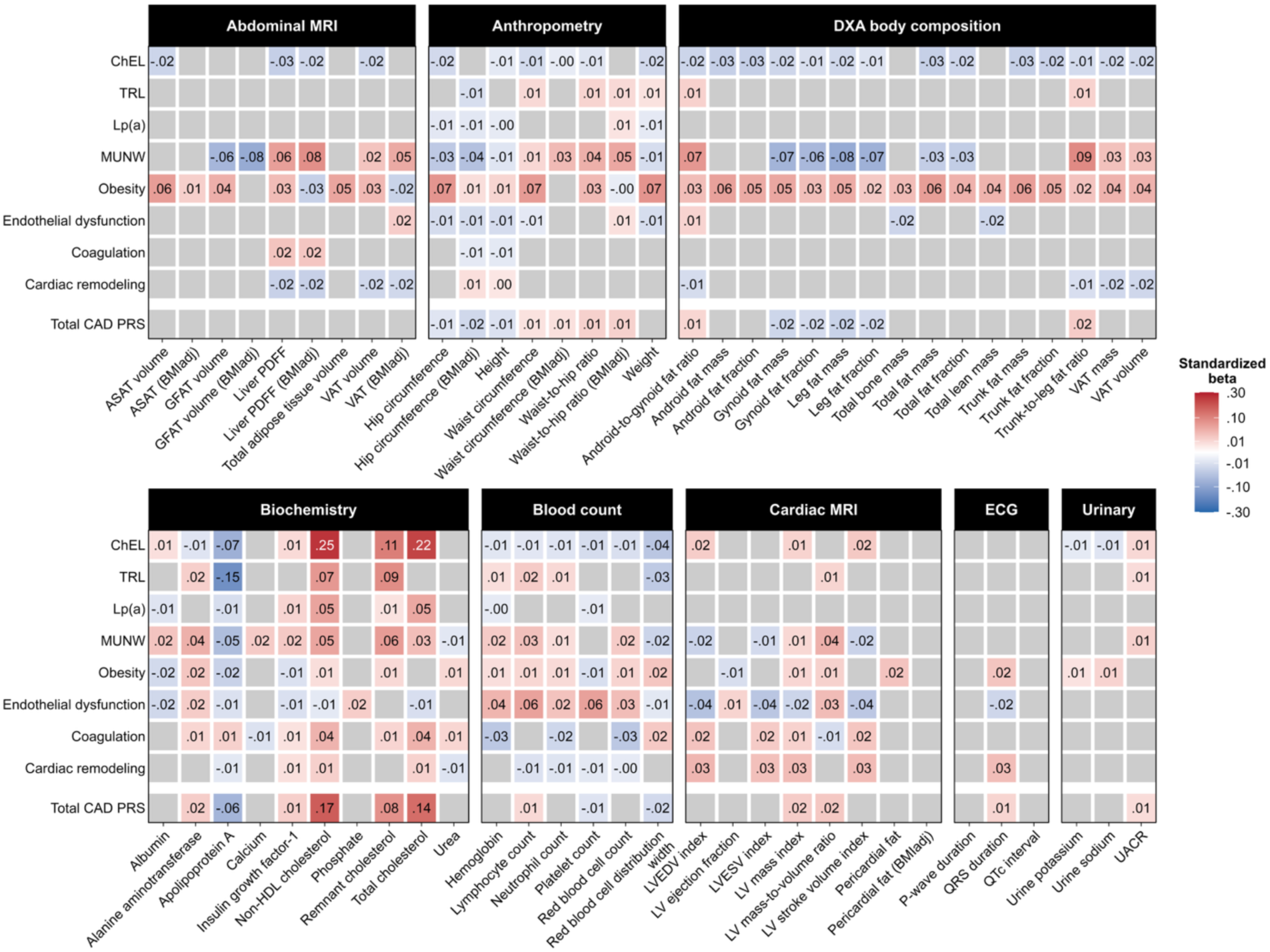
Associations of process-specific polygenic risk scores with an extended panel of clinical and imaging biomarkers. Standardized associations of each process-specific polygenic risk scores and of the total CAD PRS with quantitative traits in UKBB, grouped by measurement domain: abdominal MRI, anthropometry, DXA body composition, biochemistry, blood count, cardiac magnetic resonance imaging (MRI), electrocardiography and urinary markers. Estimates are standardized β-coefficients from linear regression, adjusted for age, sex and the first ten genetic principal components, with each outcome and each score standardized before analysis. Color denotes the direction and magnitude of the association (red, positive; blue, inverse), scaled as the square root of the absolute coefficient to improve resolution of smaller effects; the coefficient itself is printed in each cell. Grey cells indicate associations that did not reach statistical significance after correction for multiple comparisons across all tested markers (P_Bonferroni_<0.05). ASAT, abdominal subcutaneous adipose tissue; BMIadj, adjusted for body mass index; ChEL, cholesterol-enriched lipoproteins; DXA, dual-energy X-ray absorptiometry; FM, fat mass; GFAT, gluteofemoral adipose tissue; Lp(a), lipoprotein(a); LV, left ventricular; LVEDV, left ventricular end-diastolic volume; LVESV, left ventricular end-systolic volume; MUNW, metabolically-unhealthy normal weight; PDFF, proton density fat fraction; TRL, triglyceride-rich lipoproteins; UACR, urine albumin-to-creatinine ratio; UKBB, UK Biobank; VAT, visceral adipose tissue.

**Extended Data Figure 5.**
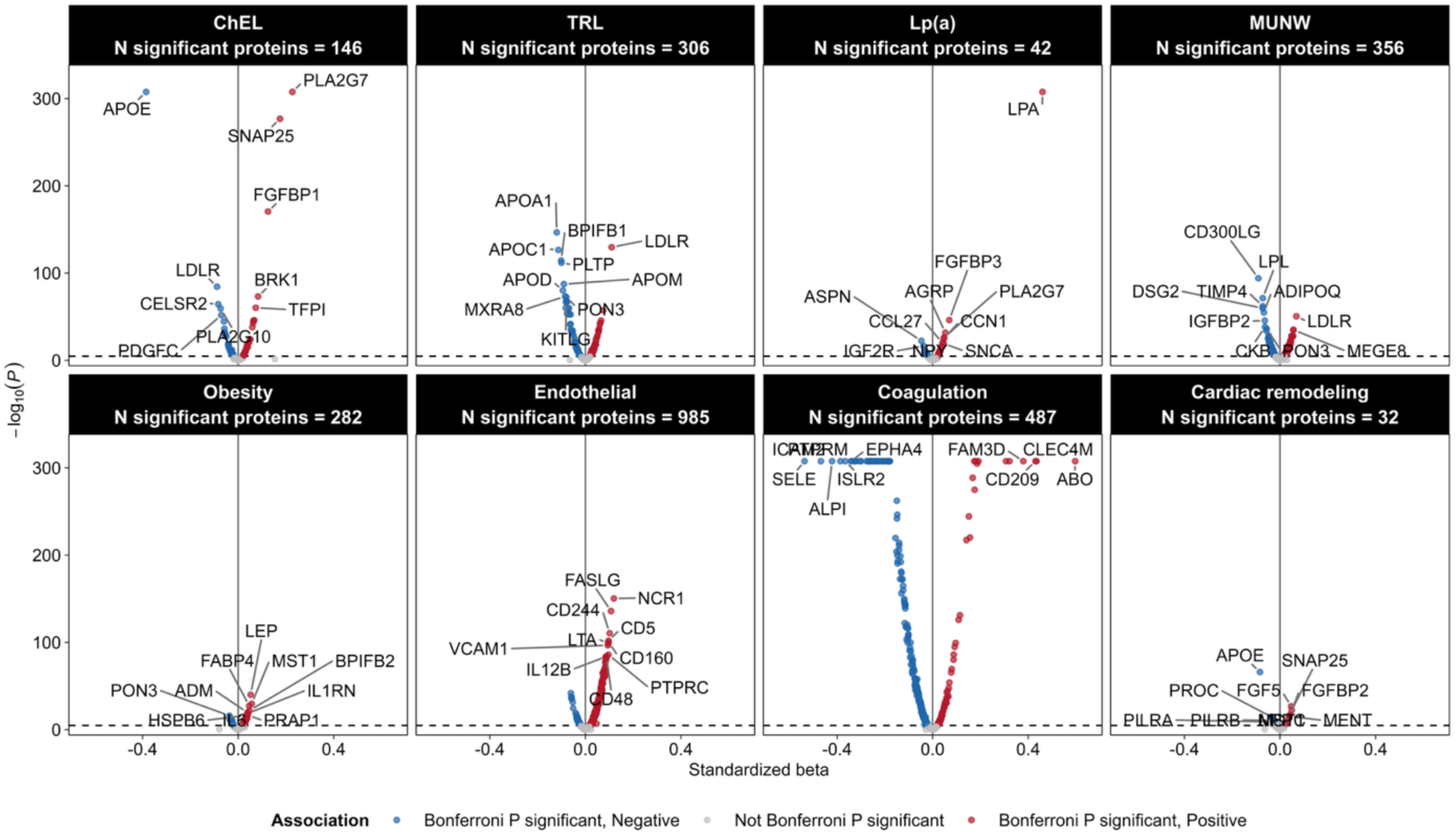
Proteome-wide associations of the process-specific polygenic risk scores. Associations of each pPRS with 2,923 plasma proteins measured by Olink in UK Biobank, shown as volcano plots of the standardized β-coefficient against −log_10_(P). Estimates are from linear regression adjusted for age, sex and the first ten genetic principal components, with each protein and each score standardized before analysis. Points are colored by direction of association (red, positive; blue, inverse) among proteins reaching statistical significance after Bonferroni correction; grey points did not reach significance, and the dashed horizontal line marks the Bonferroni-significance threshold. The number of significantly associated proteins is given above each panel. The ten proteins with the smallest P values are labelled in each panel, identified by gene symbol. Protein abbreviations are explained in **Supplementary Table 12.** ChEL, cholesterol-enriched lipoproteins; TRL, triglyceride-rich lipoproteins; Lp(a), lipoprotein(a); MUNW, metabolically unhealthy normal weight.

**Extended Data Figure 6.**
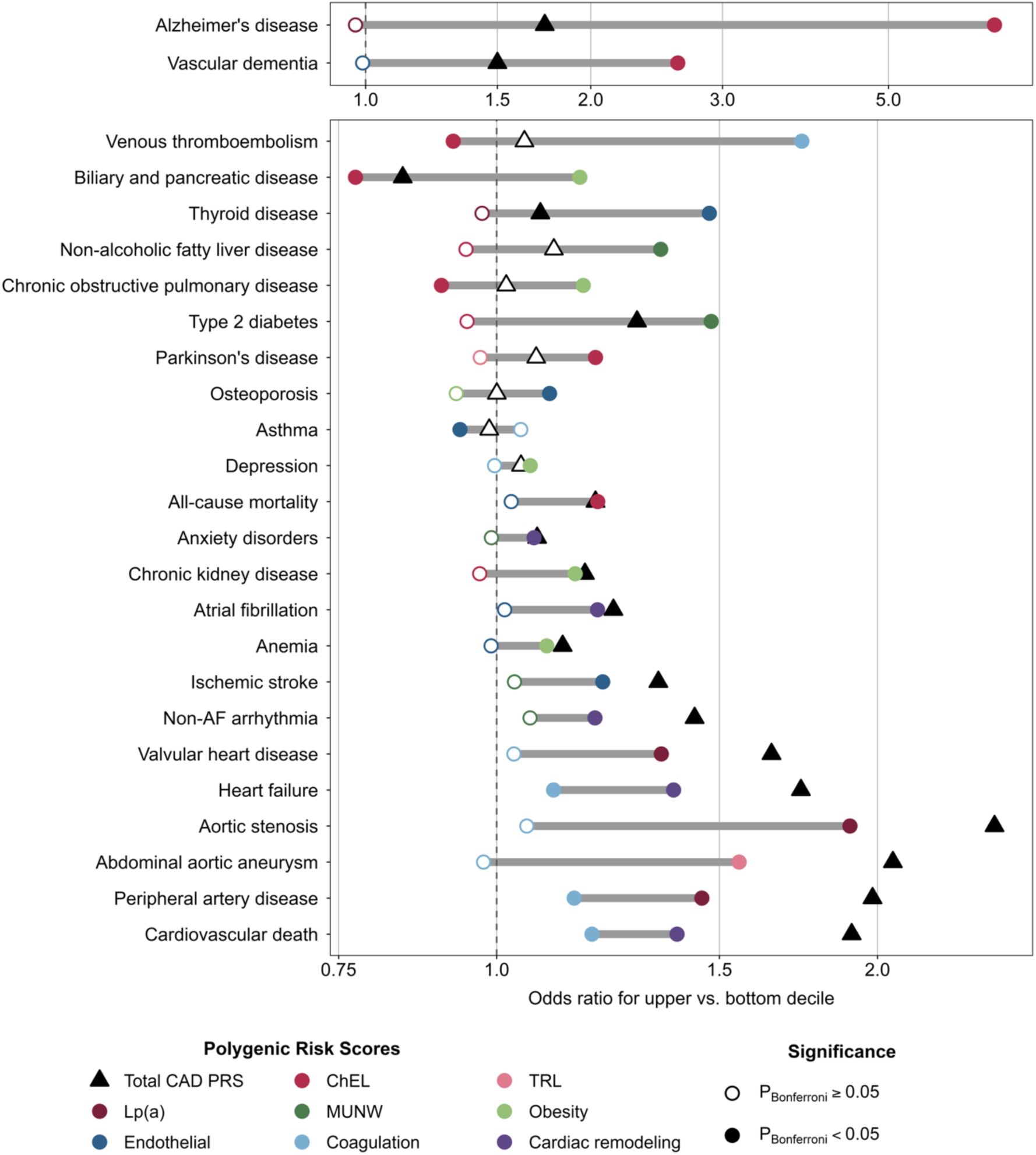
Range of associations of the process-specific polygenic risk scores with comorbidities and mortality, compared with the total CAD polygenic risk score. Odds ratios for the top compared with the bottom decile of each score for comorbidities and mortality in UK Biobank, shown for outcomes with at least one Bonferroni-significant association with a process-specific polygenic risk score (pPRS). For each outcome, circles mark the pPRSs with the highest and lowest odds ratios, connected by a bar, and triangles mark the total CAD polygenic risk score. Filled symbols indicate P<0.05 after Bonferroni correction within each score across 32 outcomes. Models were adjusted for age, sex and the first 10 genetic principal components. CAD, coronary artery disease; ChEL, cholesterol-enriched lipoproteins; Lp(a), lipoprotein(a); MUNW, metabolically unhealthy normal weight; TRL, triglyceride-rich lipoproteins.

**Extended Data Figure 7.**
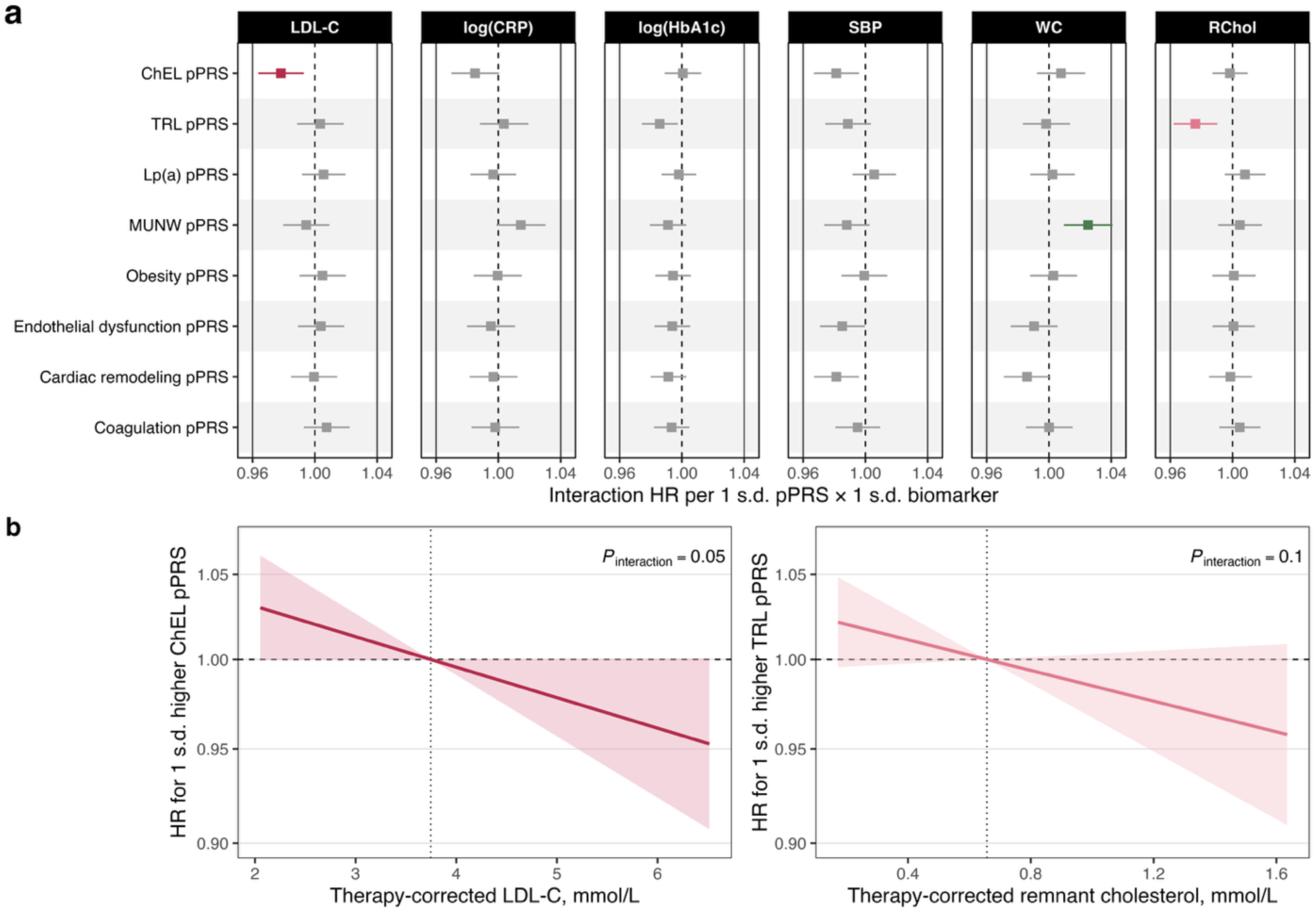
Interactions between the process-specific polygenic risk scores and conventional cardiovascular risk factors in relation to incident myocardial infarction. (a) Hazard ratios for the interaction between each pPRS and each conventional risk factor in relation to incident myocardial infarction, expressed per 1-s.d. higher pPRS and per 1-s.d. higher biomarker, in UKBB participants free of atherosclerotic cardiovascular disease at baseline. Estimates are from Cox proportional hazards models adjusted for age, sex and the first ten genetic principal components, each fitted with the pPRS, the biomarker and their product term. Colored symbols denote interactions reaching Bonferroni-corrected significance and grey symbols those that did not. (b) Hazard ratio for myocardial infarction per 1-s.d. higher ChEL pPRS across the range of therapy-corrected LDL cholesterol (left) and per 1-s.d. higher TRL pPRS across the range of therapy-corrected remnant cholesterol (right), after additional adjustment for body mass index, smoking, remaining risk factors and estimated glomerular filtration rate. Shaded areas indicate 95% confidence intervals and the dashed horizontal line the null. *P*_interaction_ values are from likelihood ratio tests of the product term. ChEL, cholesterol-enriched lipoproteins; TRL, triglyceride-rich lipoproteins; UKBB, UK Biobank.

